# Vagus Nerve Stimulation in Intracerebral and Subarachnoid Haemorrhage: A Systematic Review, Narrative Synthesis and Exploratory Meta-Analysis

**DOI:** 10.64898/2026.09.14.26362993

**Authors:** Matthew Tupper, Matthew Myers, Sheharyar Baig, Mudasar Aziz, Joyce Balami, Li Su, Arshad Majid, Ali N Ali

## Abstract

**Background/Objectives:** Haemorrhagic stroke, comprising intracerebral haemorrhage (ICH) and subarachnoid haemorrhage (SAH), accounts for a disproportionate share of stroke mortality and disability, and its incidence rises steeply with age. Secondary injury processes, including neuroinflammation, cerebral oedema, autonomic dysfunction and vasospasm, are potentially modifiable. Vagus nerve stimulation (VNS) engages several of these pathways, but its therapeutic role remains uncertain. We reviewed the preclinical and clinical evidence for VNS in haemorrhagic stroke.

**Methods:** PubMed and MEDLINE were searched from inception to 1 June 2026 for preclinical studies and randomised clinical trials of VNS in ICH or SAH. Evidence was synthesised narratively; quality was assessed using SYRCLE and PEDro. Comparable binary functional outcomes (modified Rankin Scale [mRS] 0–2) were pooled using a random-effects inverse-variance model.

**Results:** Nine reports were included: four preclinical (179 animals) and five clinical (166 participants representing four independent cohorts). Preclinical studies reported reduced inflammatory signalling, smaller haematoma volume, lower aneurysm rupture rates, improved survival and enhanced motor recovery, with attenuated effects in severe injury models. Clinically, non-invasive VNS was feasible and well tolerated, with signals of reduced tumour necrosis factor alpha and interleukin-6, less radiographic vasospasm and delayed cerebral ischaemia, and increased parasympathetic activity. Pooled favourable functional outcome favoured VNS but was not statistically significant (odds ratio [OR] 1.82, 95% confidence interval [CI] 0.68–4.86; p = 0.24; I² = 0%). Excluding major haemorrhage yielded OR 2.85 (95% CI 0.97– 8.36; p = 0.06).

**Conclusions:** VNS is biologically active and feasible after haemorrhagic stroke but remains unproven. Adequately powered, severity-stratified multicentre trials with standardised stimulation protocols and age-appropriate outcomes are required.

## 1. Introduction

### 1.1. Haemorrhagic Stroke in an Ageing Population

Haemorrhagic stroke encompasses intracerebral haemorrhage (ICH), bleeding directly into the brain parenchyma, and subarachnoid haemorrhage (SAH), bleeding into the subarachnoid space surrounding the brain. Together they account for approximately 10–15% of all strokes, yet they carry a disproportionate burden of harm, contributing around 40% of all stroke deaths and approximately half of the healthy life-years lost to stroke globally [1]. Despite advances in neurocritical care, including blood pressure management, haemostatic therapy and early aneurysm securing, outcomes after ICH and SAH remain poor [2]. Current management is largely supportive and directed at limiting further injury rather than actively promoting neurological recovery [3].

Both conditions are strongly age-related. The incidence of ICH rises steeply beyond the sixth decade, and rising rates of oral anticoagulant use, the accumulating prevalence of cerebral amyloid angiopathy and demographic ageing are expected to increase absolute case numbers over the coming decades. Older adults presenting with haemorrhagic stroke more frequently have pre-existing frailty, multimorbidity, cognitive impairment and polypharmacy, and consistently experience higher mortality, greater dependency and more limited access to rehabilitation than younger patients [2]. Age-related autonomic changes, including reduced resting vagal tone and baroreflex sensitivity, together with the low-grade systemic inflammatory state often described as inflammaging, may plausibly alter both the pathophysiology of secondary injury and the response to interventions that act through autonomic and immune pathways. Interventions capable of attenuating secondary injury without surgical risk are therefore of particular relevance to the older populations who bear most of this disease burden.

### 1.2. Secondary Neurological Injury Mechanisms

Secondary injury processes are complex, interconnected and evolve over hours to days following the initial haemorrhage [4]. In ICH, haematoma expansion may occur during the acute phase, increasing compression of surrounding tissue and worsening neurological injury [5]. The accumulation of blood and its breakdown products within the parenchyma initiates an inflammatory response involving activation of microglia and astrocytes and infiltration of peripheral immune cells. Subsequent release of pro-inflammatory cytokines contributes to blood–brain barrier (BBB) disruption, permitting further infiltration of inflammatory mediators and amplifying tissue damage [6]. BBB dysfunction also promotes cerebral oedema, which raises intracranial pressure, impairs cerebral perfusion and exacerbates perihaematomal injury [7].

Although many of these mechanisms also follow SAH, additional processes contribute to poor outcomes. Cerebral vasospasm, characterised by delayed narrowing of the cerebral arteries, commonly develops several days after aneurysmal rupture and may substantially reduce cerebral blood flow. This can precipitate delayed cerebral ischaemia (DCI), a major cause of neurological deterioration and long-term disability among SAH survivors, although DCI is now recognised to arise from a broader set of mechanisms than large-vessel vasospasm alone, including microcirculatory dysfunction, cortical spreading depolarisation and microthrombosis [8]. Together, these processes extend injury well beyond the initial bleed and represent the principal modifiable therapeutic target following haemorrhagic stroke. They are summarised in Figure 1a.

**Figure 1.**
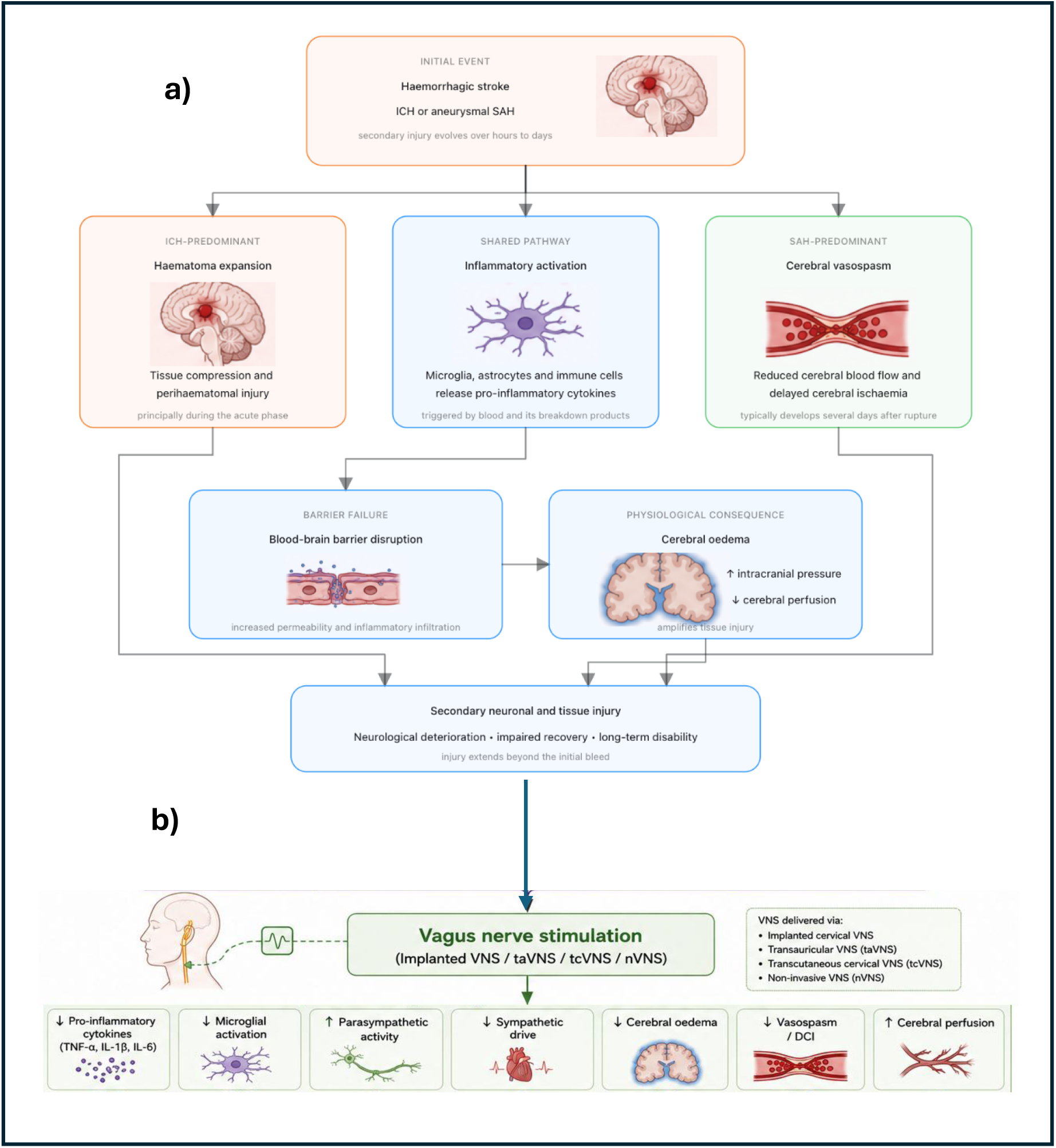
(a) Principal secondary brain injury processes following intracerebral haemorrhage and aneurysmal subarachnoid haemorrhage. (b) Proposed mechanisms through which vagus nerve stimulation may attenuate secondary brain injury and the relationship between biological target engagement and functional recovery.

### 1.3. Vagus Nerve Stimulation

Vagus nerve stimulation (VNS) has emerged as a candidate therapeutic strategy capable of targeting several of the pathological mechanisms implicated in secondary brain injury. The vagus nerve is the tenth cranial nerve and serves as a major bidirectional communication pathway between the brain and peripheral organs, with important roles in autonomic regulation, inflammatory control and physiological homeostasis [9]. Traditionally, VNS has been delivered via surgically implanted electrodes wrapped around the cervical vagus nerve and connected to a pulse generator implanted in the chest wall. More recently, non-invasive techniques have been developed, including transcutaneous auricular VNS (taVNS) and non-invasive transcutaneous cervical VNS (tcVNS), which avoid the risks of surgical implantation while improving accessibility and ease of bedside delivery [10].

VNS is an established treatment for drug-resistant epilepsy [11] and is also approved for treatment-resistant depression and certain primary headache disorders [12]. Its established safety profile and the growing understanding of its biological mechanisms have driven increasing interest in its application across a range of neurological disorders, including traumatic brain injury, disorders of consciousness, ischaemic stroke and, more recently, haemorrhagic stroke [13].

Several mechanisms have been proposed to explain its therapeutic potential. Activation of vagal pathways can suppress systemic and central inflammation through the cholinergic anti-inflammatory pathway, modulate autonomic dysfunction, influence cerebral blood flow and promote neuroplasticity [14]. Because many of these pathways overlap with the mechanisms underpinning secondary injury after ICH and SAH, VNS represents an attractive candidate capable of targeting multiple pathological processes simultaneously rather than a single downstream complication. Across experimental models of ICH and SAH, VNS has reduced neuroinflammation, attenuated cerebral oedema and improved neurological recovery [15,16]. In clinical studies, VNS has shown promise in modulating autonomic dysfunction following SAH by restoring parasympathetic activity and improving cardiovascular regulation [17], and there is emerging evidence that it may reduce the incidence and severity of cerebral vasospasm after aneurysmal SAH (aSAH) [18].

### 1.4. Rationale, Hypotheses and Objectives

The application of VNS in ICH and SAH remains in its infancy, but the evidence base is expanding rapidly and has not previously been synthesised systematically. Existing reviews of VNS in stroke have focused predominantly on ischaemic stroke and upper limb rehabilitation, with haemorrhagic subtypes either excluded or represented by small subgroups. A dedicated synthesis is therefore warranted.

We hypothesised that VNS would demonstrate beneficial effects across preclinical and clinical studies through anti-inflammatory, autonomic and neurorestorative mechanisms; that these effects would be detectable as reductions in neuroinflammation, cerebral oedema and vasospasm; and that such biological effects would be accompanied by improved neurological outcomes. We further hypothesised, on the basis of injury-severity gradients described in experimental work, that any therapeutic effect may be attenuated once primary tissue destruction became extensive.

The objectives of this review were: (i) to identify and synthesise systematically the current evidence investigating VNS in haemorrhagic stroke and related haemorrhagic brain injury; (ii) to evaluate the effects of VNS on key secondary injury mechanisms, including neuroinflammation, cerebral oedema, cerebral vasospasm and autonomic dysfunction; (iii) to assess the impact of VNS on neurological recovery, functional outcomes and disease-related biomarkers; (iv) to evaluate the safety and feasibility of VNS across preclinical and clinical studies; and (v) to compare preclinical and clinical findings and identify limitations and knowledge gaps to inform future research.

## 2. Materials and Methods

### 2.1. Design and Reporting

This systematic review was conducted and reported in accordance with the Preferred Reporting Items for Systematic Reviews and Meta-Analyses (PRISMA) 2020 statement [19] (Supplementary Table S1). Given the small and emerging evidence base, both preclinical and clinical studies were included to provide a translational overview of the therapeutic potential of VNS from experimental models through to human application. Because substantial heterogeneity was anticipated in study populations, stimulation modalities and outcome measures, a narrative synthesis was adopted, with quantitative pooling undertaken only where sufficiently comparable data were available. Preclinical and clinical evidence were extracted, appraised and synthesised separately throughout. This review was not registered with PROSPERO.

### 2.2. Eligibility Criteria

Studies were eligible if they investigated VNS in haemorrhagic stroke or related haemorrhagic brain injury and reported outcomes relating to neurological recovery, inflammation, autonomic regulation, cerebral oedema, vasospasm or functional performance. Preclinical studies using animal models of haemorrhagic injury were eligible. Clinical studies were eligible only if they employed a randomised controlled design. Studies were required to be peer-reviewed and published in English. Reviews, conference abstracts, editorials, protocols and opinion papers were excluded, as were studies that did not investigate VNS as the primary intervention or were unrelated to the review question. Although review articles were excluded from formal analysis, they were used to provide contextual background and their reference lists were screened for additional eligible reports.

### 2.3. Search Strategy

Literature searches were conducted using OVID MEDLINE with search strategies were developed to capture both haemorrhagic stroke pathologies and VNS interventions. Population terms included intracerebral haemorrhage, intracranial haemorrhage, subarachnoid haemorrhage, haemorrhagic stroke, cerebral aneurysm and aneurysmal SAH; UK and US spellings were incorporated to maximise sensitivity. Intervention terms included vagus nerve stimulation, transcutaneous VNS, transcutaneous auricular VNS, non-invasive VNS, cervical VNS, invasive VNS and associated abbreviations. Boolean operators were used to combine terms, and deep brain stimulation and transcranial magnetic stimulation were excluded using the NOT operator. The full search strategy is provided in Supplementary Table S2. Searches were conducted from database inception until 1 June 2026.

### 2.4. Study Selection

Search results were exported and duplicate records removed before screening. Titles and abstracts were assessed against the predefined eligibility criteria to remove clearly irrelevant records. Full texts were obtained and reviewed for potentially eligible reports and eligibility was reassessed using the same criteria independently by two of the authors (MT and AA). Reports were excluded if they did not investigate haemorrhagic pathology, did not use VNS as the primary intervention, did not report a relevant outcome, or represented an ineligible publication type. Reference lists of relevant reviews and included reports were also examined for eligible studies not identified through the database searches.

### 2.5. Data Extraction

Data extraction was performed using a standardised spreadsheet developed for this review, enabling consistent comparison across heterogeneous studies. Extracted information included author, publication year, country, study population, sample size, study design, VNS modality, stimulation site, stimulation frequency, intervention duration, comparator group and outcome measures. Principal findings and expanded summary results were also recorded. For preclinical studies, additional data included animal species, haemorrhage model, injury severity and experimental neurological, behavioural, inflammatory, histological and structural outcomes. For clinical studies, safety, tolerability and feasibility data were extracted, including the number and rate of adverse events and serious adverse events, their reported relationship to VNS, treatment withdrawals or discontinuations and reasons for withdrawal. Adherence data included the number of planned and completed stimulation sessions, treatment-completion rates, missed sessions and reasons for non-adherence where available. Particular attention was given to stimulation parameters because of the considerable variation between studies.

### 2.6. Outcome Measures

Given the heterogeneity of the included studies, outcomes were grouped into seven domains to facilitate comparison: haemorrhage-related outcomes; inflammation; neurological and functional recovery; survival; structural or neuroimaging outcomes; safety, feasibility and adherence; and autonomic or cardiovascular function. Preclinical haemorrhage-related outcomes included haematoma volume (ml), aneurysm rupture (%), SAH severity and other measures of haemorrhage progression (e.g. haematoma expansion). Inflammatory outcomes included pro-inflammatory cytokines and inflammatory signalling proteins, together with measures of BBB dysfunction and cerebral oedema. Neurological and functional recovery was assessed using experimental neurological scores and behavioural measures, including the modified Neurological Severity Score (mNSS), rotarod performance, beam walking and task-specific motor assessments. Survival and structural or neuroimaging measures, including neuronal survival, myelin integrity, white-matter preservation and sensorimotor-network remodelling, were additionally extracted. Clinical outcomes included safety, tolerability and feasibility; circulating and cerebrospinal fluid (CSF) inflammatory biomarkers; cerebral vasospasm, DCI and cerebral infarction; heart rate, blood pressure, heart-rate variability (HRV) and related physiological measures; and the modified Rankin Scale (mRS), Barthel Index, ongoing neurological impairment, mortality and length of hospital stay.

### 2.7. Risk of Bias Assessment

Methodological quality and risk of bias were assessed separately for preclinical and clinical studies because of inherent differences in study design and experimental methodology. Preclinical studies were assessed using the Systematic Review Centre for Laboratory Animal Experimentation (SYRCLE) risk-of-bias tool [20], developed specifically for animal intervention studies, which evaluates selection, performance, detection, attrition and reporting bias and other potential sources of bias, and additionally accounts for animal-specific considerations such as random housing, random outcome assessment and blinding procedures. Clinical studies were evaluated using the Physiotherapy Evidence Database (PEDro) scale [21], an 11-item instrument assessing randomisation, allocation concealment, baseline comparability, blinding, completeness of outcome data and statistical reporting; the eligibility-criteria item is not scored, giving a maximum of 10. One reviewer undertook the primary risk-of-bias assessments (MT) and a second reviewer independently reviewed the judgements (JB), with discrepancies resolved by discussion. Risk-of-bias findings were used to contextualise the strength of the evidence and inform interpretation. For robustness, any clinical trials included in quantitative syntheses were also assessed using the Cochrane RoB 2 tool (Supplementary table 3).

### 2.8. Statistical Analysis

Preclinical and clinical evidence were synthesised separately, with preclinical findings considered first. Because of substantial differences between animal models, intervention protocols and outcome measures, preclinical evidence was synthesised narratively and was not considered suitable for quantitative pooling.

A quantitative synthesis was undertaken for clinical reports providing sufficiently comparable binary functional-outcome data. A favourable outcome was defined as mRS 0–2 and a poor outcome as mRS 3–6. The latest available functional assessment was selected from each report: first outpatient follow-up for Huguenard et al. [18], one-month follow-up for Myers et al. [22], and hospital discharge for Rebeiz et al. [23], for which no later mRS assessment was reported. All randomised participants were retained where group allocation was known; participants for whom functional-outcome data were unavailable were conservatively classified as having a poor outcome. This approach was applied consistently across reports to avoid excluding participants on the basis of post-randomisation outcome availability. Odds ratios (ORs) with 95% confidence intervals (CIs) were calculated for favourable functional outcome in the VNS group relative to sham and visualised as Forrest Plots. Log odds ratios were pooled using a random-effects inverse-variance model, with between-study variance estimated by restricted maximum-likelihood (REML). Statistical heterogeneity was evaluated using Cochran’s Q, the I² statistic and τ² [24]. Tests were two-sided with p < 0.05 considered statistically significant. Analyses were performed in IBM SPSS Statistics version 29.0.2.0 (IBM Corp., Armonk, NY, USA). Publication bias was not formally assessed because only three reports contributed to the meta-analysis; with so few reports, funnel-plot interpretation and tests of asymmetry would be underpowered and potentially misleading.

A post hoc exploratory sensitivity analysis examined whether haemorrhage severity influenced the pooled estimate. Participants classified as having major haemorrhage (Hunt and Hess grade 5) were removed from the Myers et al. dataset [22] to match the participant inclusion in the other 2 clinical studies, and the meta-analysis repeated using the same model. This analysis was accordingly treated as an exploratory sensitivity analysis rather than a formal severity-based subgroup analysis.

## 3. Results

### 3.1. Study Selection

The database search identified 409 records and one additional record was supplied by the research team. After removal of 23 duplicates, 386 titles and abstracts were screened, and 356 records were excluded. Full texts manuscripts of 30 articles were obtained and 21 excluded because they were reviews or protocols, investigated an ineligible population or intervention, or did not report relevant outcomes. The remaining nine full texts were assessed and included, comprising five clinical reports (166 unique participants) and four preclinical reports (179 animals). Three clinical reports provided binary functional-outcome data suitable for meta-analysis. The selection process is summarised in Figure 2.

**Figure 2.**
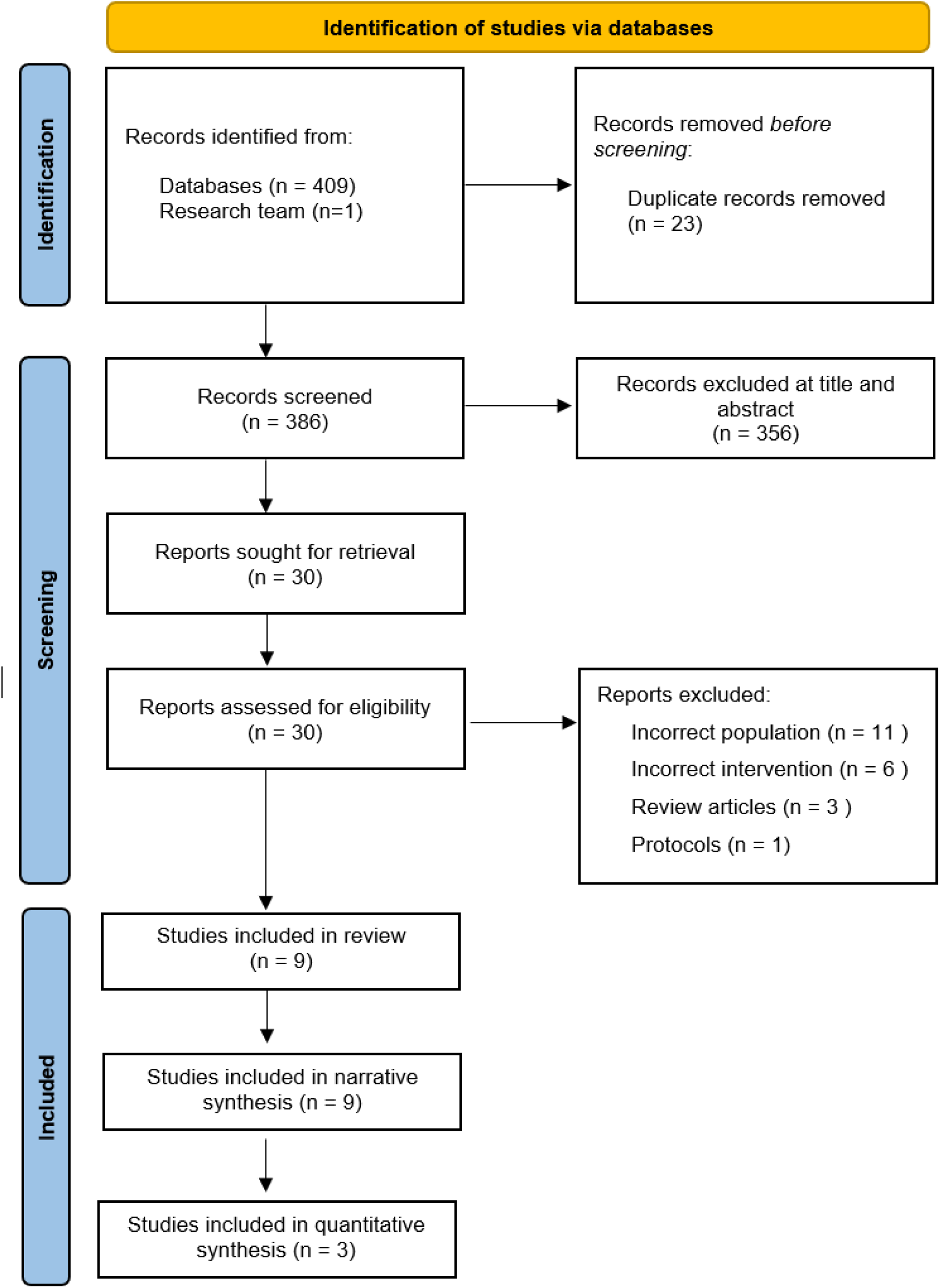
PRISMA 2020 flow diagram illustrating the identification, screening and inclusion of studies and reports.

### 3.2. Characteristics of Included Studies

Nine reports were included: five clinical and four preclinical. Three preclinical reports used rat models of ICH [16,25,26] and one used mild and severe mouse models of intracranial aneurysm rupture [27]. Preclinical sample sizes ranged from 26 to 53 animals. Interventions comprised auricular and cervical non-invasive VNS and implanted VNS paired with rehabilitation. Stimulation frequencies ranged from 25 to 30 Hz, and treatment ranged from single acute stimulation sessions to six weeks of rehabilitation-paired stimulation.

The clinical evidence comprised three published reports and one accepted manuscript involving patients with SAH or aSAH [17,18,22,23], together with one mixed acute-stroke trial that included eight participants with ICH [28]. Tan et al. [17] reported a cardiovascular analysis of 24 participants drawn from the NAVSaH trial population subsequently reported by Huguenard et al. [18]; these publications therefore do not represent independent cohorts, and the five clinical reports correspond to four independent clinical cohorts. Clinical sample sizes ranged from 24 to 71 randomised participants, although the haemorrhagic subgroup in larger Arsava et al. study comprised only eight ICH participants. Study characteristics, stimulation protocols and principal findings are presented in Table 1 (preclinical) and Table 2 (clinical).

**Table 1.** Characteristics, stimulation protocols and principal findings of included preclinical studies.

| Study, year, country | Model and species | n | VNS modality | Site | Stimulation protocol | Comparator | Outcome domains | Principal findings |
| --- | --- | --- | --- | --- | --- | --- | --- | --- |
| Zhang et al., 2025, China [16] | Collagenase ICH; rat | 50 | taVNS | Left auricular concha | 30 Hz; 30 min twice daily | Rehabilitation only | mNSS; haematoma clearance; MBP; SMI-32; FA; ALFF; corticospinal tract reconstruction; degree centrality | mNSS significantly lower at weeks 2 and 4; MBP significantly increased and SMI-32 significantly reduced at weeks 2–4; FA significantly increased; zALFF negatively correlated with mNSS ($r = -0.594$) |
| Hays et al., 2014, USA [25] | ICH; rat | 26 | Implanted (cervical) VNS paired with rehabilitation | Left cervical vagus | 30 Hz; 6-week rehabilitation programme; VNS delivered during task performance, ~15 pulses over 500 ms, 300–400 training trials per day | Rehabilitation only | Forelimb hit rate; movement speed; second-press latency; recovery percentage; lesion volume | Recovery $76.8 \pm 11.3\%$ vs $29.4 \pm 12.0\%$ ($p = 0.0062$); significant treatment effects on hit rate ($F[1,144] = 39.59$) and movement speed ($F[1,144] = 57.67$); benefit persisted after stimulation ceased; lesion volume 11.09 vs 13.55 mm<sup>3</sup> ($p = 0.38$) |
| Suzuki et al., 2019, USA/Japan [27] | Intracranial aneurysm/SAH (mild and severe models); mouse | 50 | tcVNS | Cervical | 25 Hz; two 2-min stimulations 5 min apart, once daily for 21 days | Femoral nerve stimulation | Rupture rate; SAH grade; survival; deficit-free survival; inflammatory markers; blood pressure | Mild model: rupture reduced from 80% to 29% ($p = 0.036$) and SAH grade reduced ($p = 0.025$). Severe model: rupture and SAH grade unchanged, but median survival increased from 6 to 13 days ($p = 0.003$) and deficit-free survival from 4 to 6 days ($p = 0.029$). MMP-9 reduced by 34%; reductions in IL-1β, TNF-α, CCL2, IL-6 and iNOS non-significant |
| Cáceres et al., 2025, USA/Colombia [26] | Collagenase ICH (lower- and higher-dose); rat | 53 | tcVNS | Cervical | 25 Hz; five 2-min stimulations 10 min apart, starting 30 min post-ICH | Sham (device attached, not switched on) | Haematoma volume; haemoglobin; rotarod; beam walking; Garcia score; perihematoma neuronal loss; AQP-4 | Lower-dose model: haematoma volume 5.51 vs 8.97 mm<sup>3</sup> ($p = 0.030$); rotarod decline $-29.2$ vs $-67.1$ s ($p = 0.0175$). Higher-dose model: no significant benefit (11.19 vs 12.38 mm<sup>3</sup>; $p = 0.38$). Pooled histology: greater perihematoma neuronal survival ($p = 0.0265$); AQP-4 difference borderline ($p = 0.053$) |

ALFF, amplitude of low-frequency fluctuation; AQP-4, aquaporin-4; CCL2, C-C motif chemokine ligand 2; FA, fractional anisotropy; ICH, intracerebral haemorrhage; IL, interleukin; iNOS, inducible nitric oxide synthase; MBP, myelin basic protein; MMP-9, matrix metalloproteinase-9; mNSS, modified Neurological Severity Score; tcVNS, transcutaneous cervical vagus nerve stimulation; SAH, subarachnoid haemorrhage; SMI-32, marker of non-phosphorylated neurofilament H; taVNS, transcutaneous auricular vagus nerve stimulation; TNF-α , tumour necrosis factor alpha; VNS, vagus nerve stimulation; zALFF, z-standardised ALFF.

**Table 2.**
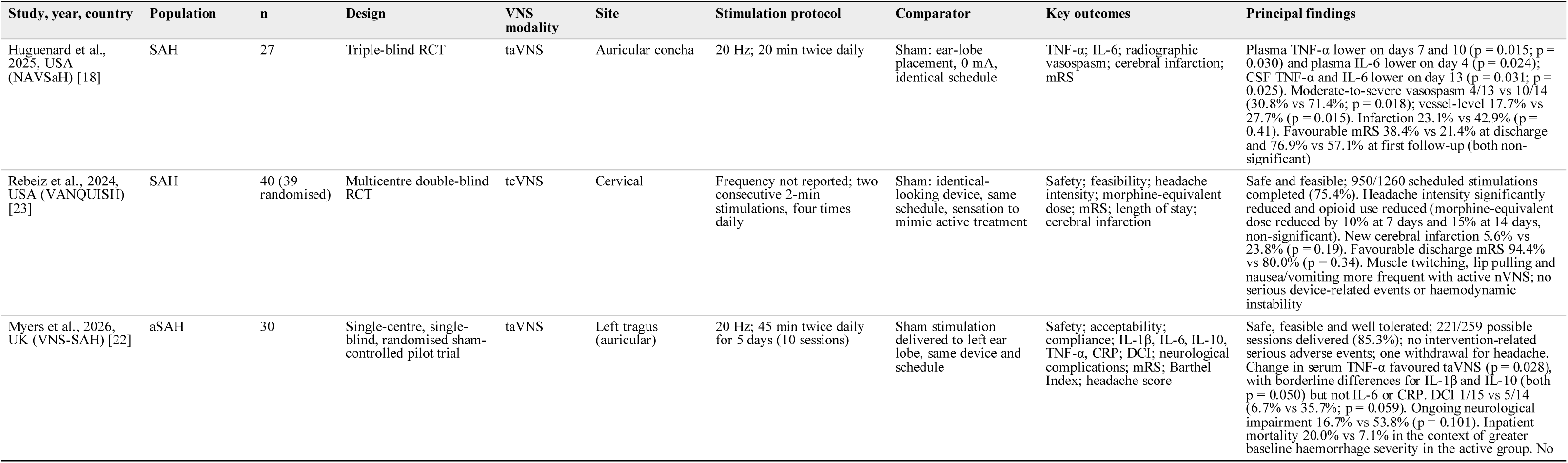
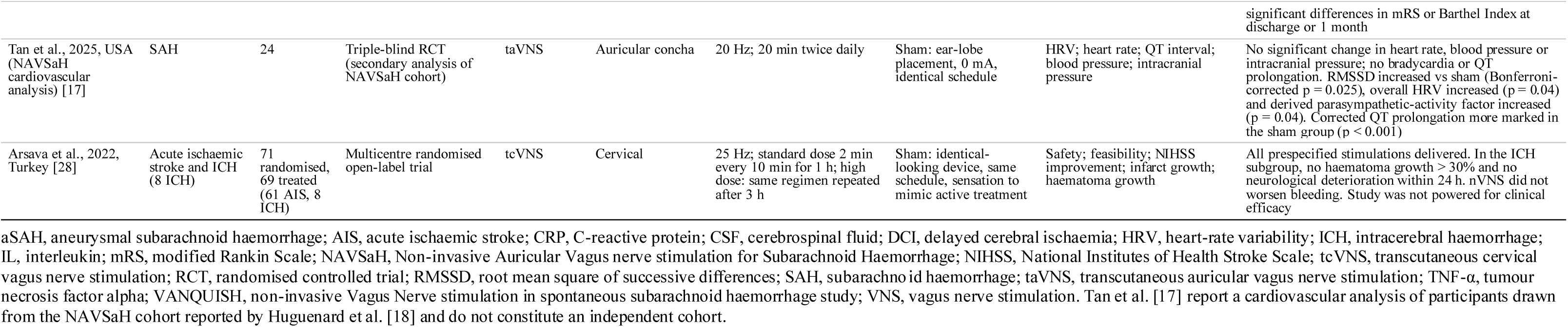
Characteristics, stimulation protocols and principal findings of included clinical studies.

### 3.3. Risk of Bias Assessment

#### 3.3.1. Preclinical Studies

The SYRCLE assessment identified variable methodological reporting (Table 3). Randomisation was reported by Suzuki et al. [27] and Cáceres et al. [26], while three reports described comparable baseline characteristics and blinded outcome assessment. All four reported their prespecified outcomes. None met the criteria for allocation concealment, blinded intervention delivery or random selection for outcome assessment.

**Table 3.** SYRCLE risk-of-bias assessment of included preclinical studies.

| SYRCLE criterion | Zhang [16] | Hays [25] | Suzuki [27] | Cáceres [26] |
| --- | --- | --- | --- | --- |
| Randomisation procedure | No | No | Yes | Yes |
| Baseline characteristics similar | No | Yes | Yes | Yes |
| Allocation concealed | No | No | No | No |
| Random animal housing | Yes | No | No | No |
| Blinding of intervention delivery | No | No | No | No |
| Random selection for outcome assessment | No | No | No | No |
| Blinding of outcome assessment | No | Yes | Yes | Yes |
| Attrition bias addressed | No | Yes | Yes | Yes |
| Reporting bias addressed | Yes | Yes | Yes | Yes |

SYRCLE, Systematic Review Centre for Laboratory Animal Experimentation. A judgement of “No” indicates that the criterion was either unmet or insufficiently reported. The “other sources of bias” domain was not classifiable from the available reports.

Attrition was a particular concern in Hays et al. [25], in which 32 of 58 animals were excluded from the principal analysis because of death, insufficient or excessive post-lesion impairment, or stimulation-device failure. Although exclusion reasons were reported and data for all animals were provided in supplementary material, the extent of exclusion introduces uncertainty regarding attrition bias. Random housing was poorly reported. A judgement of “No” indicates that a criterion was either unmet or insufficiently reported, and the “other bias” domain remained unclassified.

#### 3.3.2. Clinical Studies

PEDro scores ranged from 8 to 9 out of 10, excluding the non-scored eligibility criterion (Table 4). All five clinical reports described random allocation, concealed allocation, adequate outcome data collection, between-group comparisons and measures of variability. Four reported baseline comparability, with this criterion not recorded for the UK VNS-SAH pilot. Blinding of intervention delivery was the most frequent limitation and was not achieved in the VNS-SAH, Arsava or Tan reports. Outcome-assessor blinding was not recorded for Rebeiz et al. [23], and intention-to-treat analysis was not explicitly reported by Tan et al. [17]. These uniformly high scores should be interpreted with caution for the reasons set out in Section 4.9.

**Table 4.** PEDro assessment of the methodological quality of included clinical reports.

| PEDro criterion | Huguenard [18] | Rebeiz [23] | Myers [22] | Arsava [28] | Tan [17] |
| --- | --- | --- | --- | --- | --- |
| Eligibility criteria specified * | Yes | Yes | Yes | Yes | Yes |
| Random allocation | Yes | Yes | Yes | Yes | Yes |
| Allocation concealed | Yes | Yes | Yes | Yes | Yes |
| Baseline group similarity | Yes | Yes | No | Yes | Yes |
| Subjects blinded | Yes | Yes | Yes | Yes | Yes |
| Intervention delivery blinded | No | Yes | No | No | No |
| Assessors blinded | Yes | No | Yes | Yes | Yes |
| Outcome data ≥ 85% complete | Yes | Yes | Yes | Yes | Yes |
| Intention-to-treat analysis | Yes | Yes | Yes | Yes | No |
| Between-group statistical comparisons | Yes | Yes | Yes | Yes | Yes |
| Point estimates and variability reported | Yes | Yes | Yes | Yes | Yes |
| Total score (/10) | 9 | 9 | 8 | 9 | 8 |

PEDro, Physiotherapy Evidence Database. \* The eligibility-criteria item is not included in the total score, giving a maximum of 10. Tan et al. [17] report a secondary analysis of the cohort reported by Huguenard et al. [18].

### 3.4. Preclinical Evidence

#### 3.4.1. Haemorrhage Severity and Structural Injury

Suzuki et al. [27] induced cerebral aneurysms using stereotactic injections of elastase into the cerebrospinal fluid of mice and found that tcVNS reduced aneurysm rupture in the mild model (controlled blood pressure) from 80% with femoral-nerve stimulation to 29% (p = 0.036), and reduced SAH grade (p = 0.025). In the severe model (uncontrolled blood pressure), rupture rates and SAH grade did not differ between groups.

Cáceres et al. [26] delivered tcVNS or sham 30 minutes following induction of experimental ICH in Wistar rats, and reported a smaller mean haematoma volume following tcVNS in the lower-dose collagenase model (5.51 versus 8.97 mm³; p = 0.030), whereas no significant effect was observed in the higher-dose model (11.19 versus 12.38 mm³; p = 0.38). Haemoglobin concentration did not differ significantly at either dose. Pooled histological data across doses showed greater perihaematomal neuronal survival following tcVNS (p = 0.0265), while the difference in aquaporin-4 (AQP-4) was borderline (p = 0.053).

Zhang et al. [16] paired taVNS or sham alongside rehabilitation exercises in Sprague-Dawley rats induced with ICH (collagenase model) and reported less residual haematoma at week two with taVNS plus treadmill rehabilitation than with rehabilitation alone, although the gross anatomical comparison was based on selected samples. By week four both groups showed near-complete resolution. taVNS was also associated with greater myelin basic protein expression, lower SMI-32 expression, greater fractional anisotropy, and enhanced corticospinal-tract reconstruction when compared to sham stimulation.

Hays et al. [25] implanted cervical VNS in Sprague-Dawley ICH model rats and found no significant difference in lesion volume between VNS-paired rehabilitation and rehabilitation alone (11.09 ± 1.58 versus 13.55 ± 2.56 mm³; p = 0.38). The functional improvement associated with VNS therefore occurred without a measurable reduction in primary tissue loss.

#### 3.4.2. Neuroinflammation, Blood–Brain Barrier Disruption and Cerebral Oedema

In Suzuki et al. [27], tcVNS reduced several inflammatory mediators within the circle of Willis, but only the 34% reduction in matrix metalloproteinase-9 (MMP-9) reached statistical significance; reductions in interleukin-1β (IL-1β), tumour necrosis factor alpha (TNF-α), C-C motif chemokine ligand 2 (CCL2), interleukin-6 (IL-6) and inducible nitric oxide synthase were non-significant. Cáceres et al. [26] reported a borderline increase in AQP-4 following nVNS (p = 0.053). Neither report directly demonstrated reduced BBB permeability or cerebral oedema.

#### 3.4.3. Neurological and Motor Recovery

Neurological recovery improved in all three ICH models. Zhang et al. [16] reported lower mNSS scores with taVNS at weeks two (p < 0.05) and four (p < 0.01); increases in sensorimotor-network activity and centrality correlated with lower mNSS scores (r = −0.594, p = 0.042; r = −0.598, p = 0.040).

In Hays et al. [25], the principal analysis included 14 rats receiving VNS-paired rehabilitation and 12 receiving rehabilitation alone. Recovery of forelimb performance was greater following VNS-paired rehabilitation (76.8 ± 11.3% versus 29.4 ± 12.0%; p = 0.0062). Significant treatment effects were observed for hit rate (F[1,144] = 39.59; p = 3.54 × 10⁻⁹) and movement speed (F[1,144] = 57.67; p = 3.58 × 10⁻¹²). Training intensity did not differ between groups (p = 0.633), and the functional benefit persisted during week six after stimulation was withdrawn.

In Cáceres et al. [26], the decline in rotarod performance was smaller with tcVNS in the lower-dose model (−29.2 versus −67.1 seconds; p = 0.0175), but this benefit was not reproduced in the higher-dose model. Beam-walking and Garcia scores did not differ significantly between treatment groups.

#### 3.4.4. Survival and Physiological Outcomes

In the severe aneurysm model, median survival increased from 6 to 13 days with tcVNS (p = 0.003) and deficit-free survival increased from 4 to 6 days (p = 0.029), despite similar rupture rates and SAH grades [27]. Resting arterial blood pressure did not differ between stimulation groups.

### 3.5. Clinical Evidence

#### 3.5.1. Safety, Feasibility and Tolerability

VNS was generally deliverable without intervention-related serious adverse events. Huguenard et al. [18] reported no adverse events attributable to taVNS. In the UK VNS-SAH pilot, 221 of 259 possible sessions were delivered (85.3%) with no intervention-related serious adverse events; effects were mainly mild and transient, although one participant withdrew because of headache [22]. Rebeiz et al. [23] completed 950 of 1260 scheduled stimulations (75.4%); muscle twitching, lip pulling and nausea or vomiting were more frequent with active nVNS, but no serious device-related event or haemodynamic instability occurred. Arsava et al. [28] delivered all prespecified stimulations to 69 treated participants; among the eight participants with ICH, no haematoma growth greater than 30% and no neurological deterioration occurred within 24 hours.

#### 3.5.2. Inflammation, Vasospasm and Delayed Cerebral Ischaemia

Huguenard et al. [18] found lower plasma TNF-α in the taVNS group on days 7 and 10 (p = 0.015 and p = 0.030) and lower plasma IL-6 on day 4 (p = 0.024). CSF TNF-α and IL-6 were also lower on day 13 (p = 0.031 and p = 0.025). Moderate-to-severe radiographic vasospasm affected 4 of 13 taVNS participants and 10 of 14 sham participants (30.8% versus 71.4%; p = 0.018), and vessel-level analysis similarly favoured taVNS (17.7% versus 27.7%; p = 0.015). Cerebral infarction was numerically less frequent with taVNS, although the difference was not significant (23.1% versus 42.9%; p = 0.41).

In the VNS-SAH pilot, the change in serum TNF-α favoured taVNS (p = 0.028), with borderline between-group differences for IL-1β and interleukin-10 (both p = 0.050) but not IL-6 or C-reactive protein [22]. DCI occurred in 1 of 15 active participants and 5 of 14 sham participants (6.7% versus 35.7%; p = 0.059). Rebeiz et al. [23] also reported fewer new cerebral infarctions with nVNS than sham (5.6% versus 23.8%), although this difference was not statistically significant (p = 0.19).

#### 3.5.3. Autonomic and Cardiovascular Outcomes

In the NAVSaH cardiovascular analysis, repetitive taVNS did not significantly alter heart rate, blood pressure or intracranial pressure and did not produce bradycardia or QT prolongation [17]. Relative to sham, taVNS increased the root mean square of successive differences between heartbeats (RMSSD; Bonferroni-corrected p = 0.025), overall HRV (p = 0.04) and the derived parasympathetic-activity factor (p = 0.04). Corrected QT prolongation was more marked in the sham group (p < 0.001).

#### 3.5.4. Neurological and Functional Outcomes

Clinical functional outcomes were generally favourable in direction but statistically inconclusive. In Huguenard et al. [18], favourable mRS outcomes occurred in 38.4% versus 21.4% at discharge and 76.9% versus 57.1% at first follow-up for taVNS and sham respectively; neither between-group comparison was statistically significant. Rebeiz et al. [23] reported favourable discharge mRS outcomes in 94.4% of active participants and 80.0% of sham participants (p = 0.34), with no significant difference in length of stay, alongside significantly reduced headache intensity and reduced opioid use. In the VNS-SAH pilot, neither mRS nor Barthel Index differed significantly at discharge or one month; ongoing neurological impairment was numerically less frequent with taVNS (16.7% versus 53.8%; p = 0.101), but inpatient mortality was 20.0% with taVNS and 7.1% with sham in the context of greater baseline haemorrhage severity in the active group [22].

### 3.6. Meta-Analysis

#### 3.6.1. Included Reports and Outcome Definition

Three SAH reports contributed binary data for a favourable functional outcome: Huguenard et al. [18], Rebeiz et al. [23] and Myers et al. [22], comprising 96 participants. A random-effects inverse-variance model with REML estimation was used to pool log odds ratios.

#### 3.6.2. Primary Analysis

In the primary analysis, which included all available haemorrhage severities, the pooled estimate favoured VNS but was not statistically significant (OR 1.82, 95% CI 0.68–4.86; p = 0.24). No statistical between-study heterogeneity was detected (I² = 0%; τ² = 0.00; Q = 1.56, df = 2, p = 0.46). Individual and pooled estimates are shown in Figure 3.

**Figure 3.**
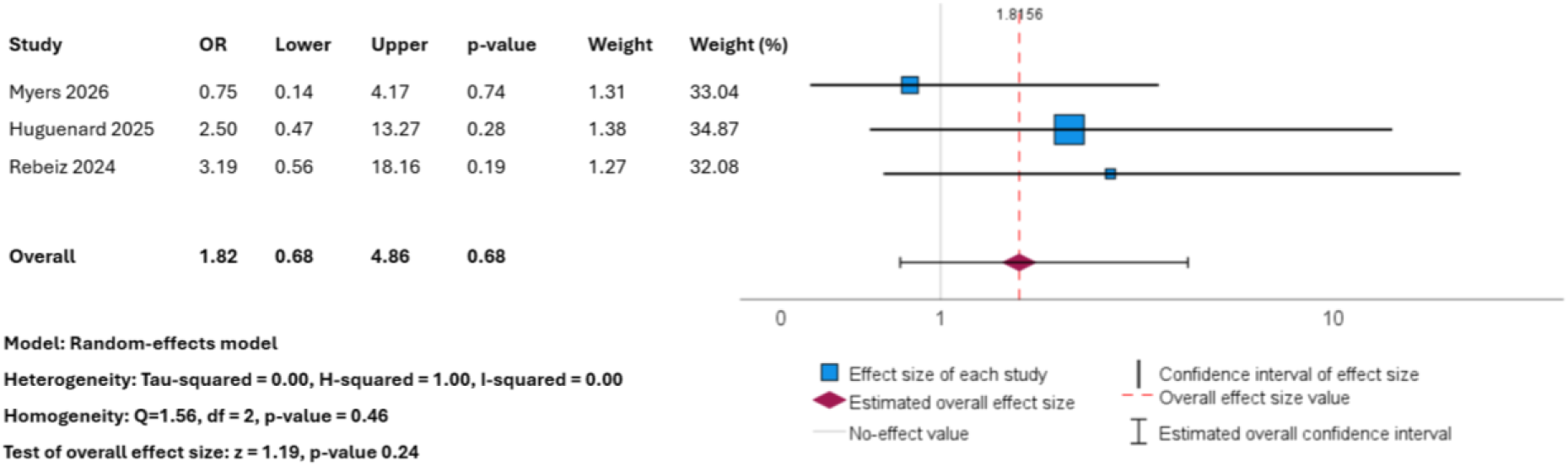
Forest plot of the association between vagus nerve stimulation and favourable functional outcome (modified Rankin Scale 0–2) in the primary random-effects meta-analysis. Odds ratios greater than 1 favour vagus nerve stimulation. CI, confidence interval; OR, odds ratio.

#### 3.6.3. Exploratory Sensitivity Analysis

In a post hoc exploratory sensitivity analysis in which participants classified as having major aSAH (Hunt and Hess grade 5, n = 4) were removed from the Myers dataset, to match the population in the other 2 studies, the pooled effect increased to OR 2.85 (95% CI 0.97–8.36; p = 0.06). No statistical heterogeneity was detected (I² = 0%; τ² = 0.00; Q = 0.04, df = 2, p = 0.98). Results are shown in Figure 4.

**Figure 4.**
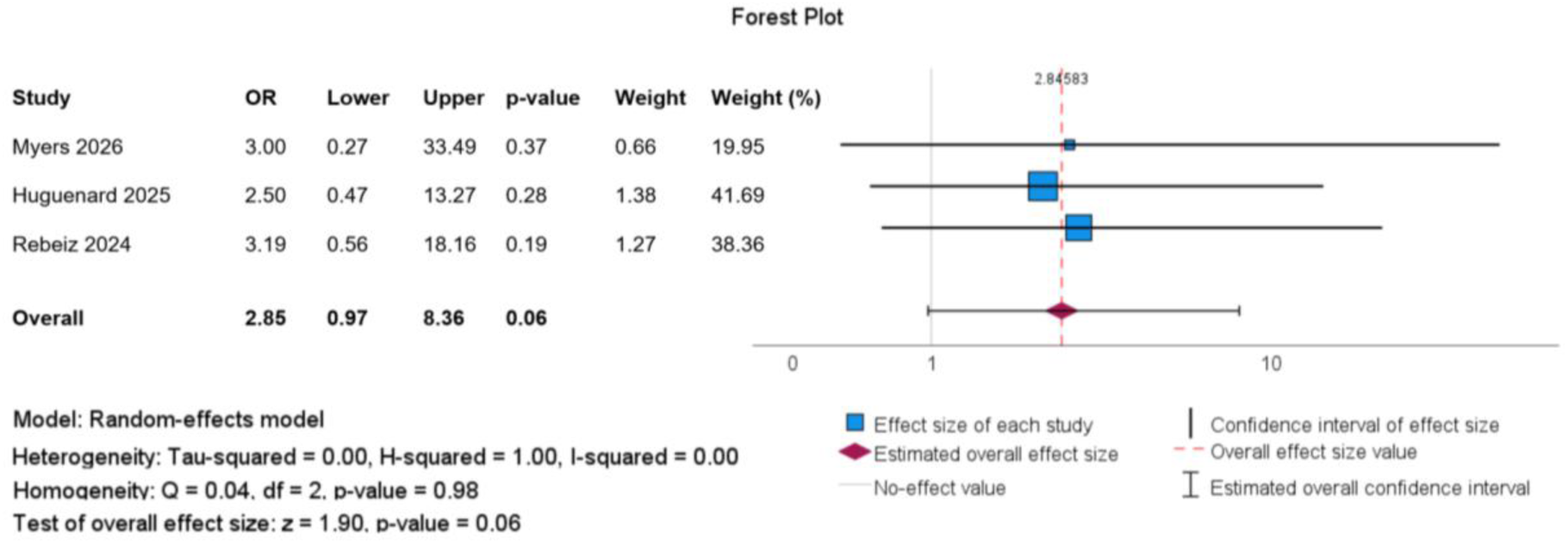
Forest plot of the post hoc sensitivity analysis excluding participants classified as having major haemorrhage (Hunt and Hess grade 5, n = 4) from the Myers dataset. The Huguenard and Rebeiz data remained unchanged. Odds ratios greater than 1 favour vagus nerve stimulation. CI, confidence interval; OR, odds ratio.

Although the sensitivity analysis produced a larger pooled effect, its confidence interval crossed the null value and the result did not reach statistical significance. Because only the Myers contribution was modified, this was not equivalent to a prospectively defined severity subgroup analysis applied consistently across all three reports. Furthermore, the small number of included reports limited the precision of both the pooled effect and the heterogeneity estimates. Individual and pooled treatment effects are summarised in Table 5.

**Table 5.** Summary of the primary and post hoc sensitivity meta-analyses of favourable functional outcome.

| Analysis | k | Pooled OR (95% CI) | p-value | Heterogeneity |
| --- | --- | --- | --- | --- |
| Primary analysis: all available haemorrhage severities | 3 | 1.82 (0.68–4.86) | 0.24 | Q = 1.56, df = 2, p = 0.46; I<sup>2</sup> = 0%; $\tau^2$ = 0.00 |
| Post hoc sensitivity analysis: major haemorrhage excluded from the Myers dataset | 3 | 2.85 (0.97–8.36) | 0.06 | Q = 0.04, df = 2, p = 0.98; I<sup>2</sup> = 0%; $\tau^2$ = 0.00 |

CI, confidence interval; df, degrees of freedom; k, number of contributing reports; OR, odds ratio. Analyses used a random-effects inverse-variance model with restricted maximum-likelihood estimation. An OR greater than 1 favours vagus nerve stimulation. Favourable outcome was defined as modified Rankin Scale 0–2. The sensitivity analysis excluded participants classified as having major haemorrhage from the Myers dataset only.

## 4. Discussion

This systematic review identified an emerging but limited evidence base indicating that VNS is biologically active following haemorrhagic brain injury. Across preclinical studies, VNS was associated with reduced haemorrhage-related injury and inflammatory signalling, alongside improvements in neurological recovery and neuroplastic remodelling, although these effects were less consistent in severe injury models. In early clinical studies, VNS was generally feasible and well tolerated in neurocritical care populations, with preliminary effects on inflammatory biomarkers, cerebral vasospasm, DCI and autonomic regulation. These favourable biological findings did not, however, consistently translate into significant improvements in disability or independence; pooled functional-outcome estimates favoured VNS but remained statistically inconclusive. Overall, the findings partially support the proposed therapeutic potential of VNS while suggesting that clinical benefit may depend on injury severity, treatment timing and stimulation protocol.

### 4.1. Biological and Translational Effects of VNS

Preclinical evidence suggests that VNS may attenuate selected components of acute brain injury, though its effects appear to depend on haemorrhage severity. Mechanistically, vagal activation is proposed to engage the cholinergic anti-inflammatory pathway, through which acetylcholine-dependent signalling at the α7 nicotinic acetylcholine receptor suppresses immune-cell production of pro-inflammatory cytokines including TNF-α, IL-1β and IL-6 [29,30]. In the mild intracranial aneurysm model, Suzuki et al. [27] reported reduced aneurysmal rupture and SAH severity following nVNS; in the severe model, rupture and SAH severity were unchanged although survival was prolonged, suggesting that VNS may influence secondary pathological consequences without preventing the primary event. Similarly, Cáceres et al. [26] observed a smaller haematoma volume and improved motor performance in the lower-dose ICH model, alongside greater perihaematomal neuronal survival overall, whereas comparable benefits were absent following more severe injury. Together these findings support possible neuroprotective and anti-inflammatory effects but suggest a biological ceiling once primary tissue injury becomes extensive. Notably, neither study directly demonstrated restoration of BBB integrity or reduced cerebral oedema, and the borderline increase in AQP-4 reported by Cáceres et al. is of uncertain direction of benefit, since AQP-4 contributes to both oedema formation and oedema clearance depending on injury phase.

Beyond effects on acute secondary injury, VNS may enhance activity-dependent neuroplasticity when paired with rehabilitation. VNS delivered in close temporal association with task practice is proposed to recruit cholinergic and noradrenergic neuromodulatory systems, strengthening plasticity within the circuits engaged during that practice and promoting task-specific cortical reorganisation [31,32]. Hays et al. [25] reported substantially greater forelimb recovery following VNS-paired rehabilitation than rehabilitation alone, with benefits persisting after stimulation was withdrawn. Because lesion volume did not differ between groups, this improvement is more plausibly explained by reorganisation of surviving neural networks than by preservation of injured tissue. Similarly, Zhang et al. [16] associated taVNS with improved neurological scores, greater myelin preservation, corticospinal-tract reconstruction and sensorimotor-network remodelling, although the observed correlations cannot establish causation. These findings are consistent with evidence from ischaemic stroke: the 108-participant VNS-REHAB trial found that implanted VNS paired with task-specific rehabilitation produced greater improvements in upper limb impairment and a higher response rate than sham-paired rehabilitation [33]. Differences in pathology, disease stage and stimulation modality nonetheless prevent direct extrapolation to ICH or SAH populations. Taken together, the preclinical evidence suggests two distinct potential roles for VNS: reduction of acute secondary injury, and later enhancement of rehabilitation-driven recovery.

Early clinical evidence provides partial support for translating preclinical anti-inflammatory findings into patients with SAH. Huguenard et al. [18] reported lower plasma and CSF concentrations of TNF-α and IL-6 at selected time points, alongside a lower incidence of moderate-to-severe radiographic vasospasm following taVNS, with numerically less frequent cerebral infarction. Myers et al. [22] found that the change in serum TNF-α favoured taVNS, with borderline findings for IL-1β and IL-10 but no significant differences in IL-6 or C-reactive protein, and DCI occurred less frequently in the active group although the result remained inconclusive. Rebeiz et al. [23] also reported numerically fewer new cerebral infarctions with nVNS. Collectively these findings are compatible with vagal modulation of inflammatory and vascular secondary injury. However, the studies did not establish that cytokine changes caused reductions in vasospasm, DCI or infarction, and reduced radiographic vasospasm should not be interpreted as direct evidence of vasodilation or improved cerebral perfusion. The findings indicate preliminary biological activity rather than established clinical efficacy.

Autonomic findings provide further evidence that taVNS engaged relevant physiological pathways in patients with SAH. In a secondary analysis of the NAVSaH cohort, Tan et al. [17] reported increased RMSSD, overall HRV and parasympathetic activity without clinically concerning changes in heart rate, blood pressure or intracranial pressure. This pattern is consistent with parasympathetic modulation and suggests that HRV could serve as a marker of physiological target engagement, although it remains an indirect and potentially confounded measure in sedated, ventilated patients receiving vasoactive and sedative drugs. Because Tan et al. analysed participants from the trial reported by Huguenard et al., these findings are complementary rather than independent confirmation. Across the Huguenard, Myers and Rebeiz reports, no intervention-related serious adverse event was identified and reported effects were predominantly mild and transient. Arsava et al. [28] also observed no substantial haematoma growth or early neurological deterioration among the eight participants with ICH. Overall, VNS appears feasible and generally well tolerated in neurocritical care settings, but small samples cannot exclude uncommon adverse events or establish safety specifically in ICH.

### 4.2. A Severity Threshold for VNS Responsiveness

Although the biological findings were encouraging, evidence that VNS improves patient-centred functional recovery was less conclusive. The meta-analysis of three SAH reports produced a pooled estimate favouring VNS for achieving a favourable mRS outcome, but the confidence interval crossed the null value. This should not be interpreted as evidence that VNS has no effect; rather, the wide interval remains compatible with clinically important benefit, with no effect, and with modest harm. No statistical heterogeneity was detected, with I² and τ² both equal to zero, but heterogeneity statistics have limited power when only three reports are available and cannot exclude clinically important differences in stimulation modality, haemorrhage severity, follow-up timing and outcome availability. The pooled result should therefore be regarded as hypothesis-generating rather than confirmatory.

The discrepancy between favourable biological signals and inconclusive functional outcomes may reflect inadequate statistical power, but it may also indicate that treatment response varies according to the severity of the primary haemorrhagic injury. The post hoc sensitivity analysis explored this possibility. After participants classified as having major haemorrhage were removed from the Myers dataset, the estimated benefit of VNS increased and moved closer to statistical significance, though the confidence interval still crossed the null. This is directionally consistent with the preclinical findings of Suzuki et al. [27] and Cáceres et al. [26], in which benefits were less pronounced in severe haemorrhage models. One explanation is that VNS primarily modifies secondary inflammatory, vascular and neuroplastic processes and therefore has limited capacity to influence outcome once extensive irreversible primary tissue destruction has occurred. By analogy with the ischaemic penumbra, VNS may require a sufficient volume of salvageable, structurally intact tissue for its effects to be expressed in functional terms.

We propose this as a formal, falsifiable hypothesis: that the effect of VNS on functional outcome after haemorrhagic stroke is modified by baseline injury severity, such that treatment effect declines as the volume of irreversibly injured tissue increases. This hypothesis is testable through stratified randomisation and a prespecified treatment-by-severity interaction test, and it carries direct design implications, since trials enrolling unselected populations including a substantial proportion of high-grade haemorrhage may dilute a genuine effect in those most likely to respond. It must be emphasised that the present sensitivity analysis cannot establish this: only the Myers contribution was altered so that participants were matched across clinical aSAH studies. The analysis was neither prespecified nor tested for a formal interaction. The apparent increase could reflect changes in event distribution or chance alone.

### 4.3. Haemostatic Effects and the “Neural Tourniquet”: An Untested Hypothesis in Haemorrhagic Stroke

One mechanism with specific and previously unexamined relevance to haemorrhagic stroke is the vagal regulation of haemostasis. Electrical VNS has been shown to prime circulating platelets through acetylcholine-secreting choline acetyltransferase-positive T lymphocytes in the spleen acting at platelet α7 nicotinic acetylcholine receptors, increasing intracellular calcium and alpha-granule release and accelerating local thrombin generation at sites of tissue injury without inducing systemic thrombosis. This pathway, termed the neural tourniquet, reduces blood loss in multiple experimental haemorrhage models and can bypass factor VIII deficiency in murine haemophilia A. Two observations in the present review are at least consistent with such an effect: the reduced aneurysm rupture rate and SAH grade reported by Suzuki et al. [27] in the mild model, and the smaller haematoma volume reported by Cáceres et al. [26] in the lower-dose model, in which nVNS was initiated 30 minutes after collagenase injection, during the window in which haematoma expansion occurs.

This generates a distinct hypothesis that has not, to our knowledge, been tested clinically: that acutely administered VNS may reduce haematoma expansion after ICH. Haematoma expansion is one of the few modifiable determinants of outcome in ICH and remains an attractive therapeutic target. If VNS exerts a pro-haemostatic effect at the site of injury without systemic prothrombotic consequences, it would be mechanistically distinct from, and potentially complementary to, blood pressure lowering and haemostatic agents. The reassuring absence of haematoma growth exceeding 30% among the eight ICH participants in Arsava et al. [28] is compatible with this hypothesis but is far too small to test it. This mechanism also has a corollary relevant to SAH: any pro-haemostatic effect would need to be balanced against the ischaemic component of DCI and against the elevated risk of venous thromboembolism in immobilised neurocritical care patients. Future haemorrhagic stroke trials should therefore incorporate serial imaging to quantify haematoma expansion, alongside systematic ascertainment of thrombotic events, so that both potential benefit and potential harm from this pathway can be evaluated directly.

### 4.4. Biological Target Engagement Versus Functional Recovery

The findings reveal an important distinction between biological activity and clinically meaningful functional recovery. VNS produced favourable signals involving inflammatory cytokines, radiographic vasospasm and autonomic regulation, suggesting engagement of relevant physiological pathways. Improvements in mRS, Barthel Index and other measures of disability were nonetheless statistically inconclusive. This is not necessarily contradictory: biomarkers represent intermediate points within a complex causal pathway, and modest biological changes may be insufficient to overcome extensive primary tissue destruction, DCI, hydrocephalus, systemic complications or differences in rehabilitation exposure.

The history of vasospasm-directed therapy in aSAH provides a cautionary precedent. Interventions that convincingly reduced angiographic vasospasm have repeatedly failed to improve functional outcome [34], reflecting the now well-established dissociation between large-vessel narrowing and DCI, which also arises from microcirculatory dysfunction, cortical spreading depolarisation, microthrombosis and early brain injury [8]. The reduction in radiographic vasospasm reported by Huguenard et al. [18] should therefore be regarded as a mechanistic signal rather than a surrogate for benefit, and future trials of VNS in aSAH should be powered for patient-centred functional endpoints rather than for radiographic vasospasm.

Functional benefit may also have been underestimated by the available outcome assessments. The mRS provides a broad measure of global disability and may not detect more specific changes in motor, cognitive or autonomic function, while dichotomisation into favourable and poor outcomes discards information; ordinal shift analysis would be more efficient in trials of this size. Follow-up periods may also have been too short to identify delayed recovery. Most clinical studies were designed primarily to assess safety, feasibility or biological effects rather than being powered for functional efficacy. The current evidence therefore supports physiological target engagement but does not establish clinical effectiveness. The proposed relationships between VNS, biological target engagement and functional recovery are summarised in Figure 1b.

### 4.5. Immunomodulation, Infection Risk and the Spleen

A consequence of the proposed mechanism that has received little attention in this literature is that the cholinergic anti-inflammatory pathway is, by design, immunosuppressive. Acute haemorrhagic stroke already induces a state of stroke-induced immunodepression [35], and infection, particularly pneumonia, is among the commonest complications and a strong determinant of poor outcome, especially in older and dysphagic patients [36]. It is therefore biologically plausible that effective vagal suppression of pro-inflammatory cytokine production could increase susceptibility to hospital-acquired infection, and equally plausible that any such effect might offset neurological benefit. None of the included clinical reports was designed or powered to detect this, and infection outcomes were not systematically reported in the studies synthesised here. This is an important evidential gap: hospital-acquired infection should be a prespecified safety outcome in all future trials of VNS in acute haemorrhagic stroke, reported with the same rigour as device-related adverse events. The same reasoning implies that splenic integrity may modify treatment response, since the spleen is the principal effector organ of this pathway; splenectomy and functional hyposplenism are readily ascertainable and should be recorded.

### 4.6. Considerations Specific to Older Adults

Haemorrhagic stroke is predominantly a disease of later life, yet none of the included studies reported age-stratified results, and the preclinical work was conducted exclusively in young adult animals. This is a substantive translational gap, since several age-related changes could plausibly modify the effect of VNS in either direction. Resting vagal tone and baroreflex sensitivity decline with age [37], and HRV falls correspondingly, which may reduce the physiological headroom available for parasympathetic augmentation and complicate the use of HRV as a target-engagement biomarker in older cohorts [38]. Conversely, the chronic low-grade inflammatory state characteristic of ageing might render older patients more, rather than less, responsive to an anti-inflammatory intervention. Anticholinergic burden is high in older inpatients [39] and could pharmacologically antagonise a pathway that depends on nicotinic acetylcholine receptor signalling; beta-blockers, commonly prescribed in this population, alter HRV independently of vagal stimulation. A useful precedent exists in the remote ischaemic conditioning literature, in which sulphonylurea therapy abolishes the cardioprotective effect of conditioning [40], illustrating how routinely prescribed drugs can silently negate a conditioning-type intervention.

Practical considerations also differ. Auricular stimulation depends on skin integrity and electrode contact, both of which may be affected by age-related dermal changes, oedema and prolonged supine positioning. Adherence to twice-daily stimulation may be harder to achieve in patients with delirium or agitation, and delirium itself is a plausible, inflammation-linked outcome that VNS might influence but which no included study measured. Finally, the mRS performs poorly as an outcome measure in older adults with pre-existing disability; pre-morbid mRS was not consistently reported, and analyses that do not account for baseline function risk both misclassification and loss of power. Future trials should report age-stratified results, record pre-morbid function, anticholinergic burden and frailty, and include cognition, delirium and discharge destination among their outcomes. Encouragingly, VNS paired with rehabilitative training remains effective in 18-month-old rats after ischaemic motor cortex lesion, producing near-complete recovery of forelimb function compared with approximately one-third recovery with rehabilitation alone [41], suggesting that advanced age need not preclude benefit.

### 4.7. Clinical Implications and Future Research

Taken together, the evidence suggests that the most realistic clinical role for VNS would be as a stage-specific adjunct rather than a treatment for the primary haemorrhage. During acute neurocritical care, non-invasive auricular or cervical VNS could be delivered following aneurysm treatment or medical stabilisation to modulate secondary injury processes, particularly inflammation and autonomic dysregulation, with possible downstream effects on vasospasm and DCI. Its capacity for repeated bedside administration without surgical implantation is clinically attractive and is of particular value in frail patients for whom invasive procedures carry disproportionate risk; the studies by Huguenard et al. [18], Rebeiz et al. [23] and Tan et al. [17] provide preliminary evidence that this approach is feasible and reasonably tolerated. VNS would not, however, be expected to reverse mass effect, established infarction or extensive irreversible tissue destruction. Following medical stabilisation, a separate potential role involves pairing VNS with task-specific rehabilitation to reinforce neural circuits activated during therapy, supported by the preclinical findings of Hays et al. [25] and Zhang et al. [16] together with rehabilitation-paired VNS in ischaemic stroke [33], although this has not yet been tested clinically after haemorrhagic stroke. Acute neuroprotection and rehabilitation enhancement should therefore be treated as distinct applications requiring different stimulation protocols, outcome measures and trials.

HRV may offer one route towards personalising VNS because it can be measured non-invasively and may indicate whether stimulation has engaged the intended autonomic pathways. Tan et al. [17] observed increases in RMSSD, overall HRV and parasympathetic activity following taVNS, raising the possibility that baseline HRV or an early physiological response could identify patients demonstrating target engagement. Supporting the biological plausibility of this approach, Cooper et al. [42] found inverse associations between HRV and several systemic inflammatory markers, with low-frequency HRV inversely associated with fibrinogen, C-reactive protein and IL-6, and high-frequency HRV inversely associated with fibrinogen and C-reactive protein. However, this was a cross-sectional community study and did not establish that HRV predicts the inflammatory response to VNS or outcomes after haemorrhagic stroke. Future trials should investigate whether HRV during treatment predicts cytokine suppression, vasospasm, DCI or functional recovery, and whether stimulation parameters could safely be titrated to this response. Until such relationships are prospectively validated, HRV should be regarded as a candidate target-engagement biomarker rather than an established tool for patient selection or personalised dosing.

Future research should move beyond small, heterogeneous feasibility studies towards adequately powered, multicentre, sham-controlled trials with prospectively registered analysis plans. SAH and ICH should initially be investigated separately because their secondary injury processes, treatment pathways and relevant outcomes differ, and because the current evidence base is markedly asymmetrical: the preclinical signal is strongest in ICH, whereas almost all clinical data derive from SAH. Haemorrhage severity should be incorporated into stratified randomisation, with any treatment-by-severity analysis specified in advance. Trials should clearly distinguish between VNS administered acutely to reduce secondary injury and VNS paired with later rehabilitation. Acute trials should use a defined treatment window and assess a prespecified patient-centred functional outcome at longer-term follow-up, treating cytokines, HRV, vasospasm and DCI as secondary mechanistic or target-engagement outcomes; haematoma expansion, hospital-acquired infection and thrombotic events should be prespecified. Rehabilitation trials should pair stimulation consistently with standardised, task-specific therapy and use domain-specific recovery measures. The multicentre VNS-REHAB trial demonstrates that a blinded, standardised rehabilitation-paired design is feasible, though its efficacy findings cannot be extrapolated directly to haemorrhagic stroke [33]. Dose-finding remains an unaddressed priority: every included clinical study used a fixed protocol, and no study compared stimulation intensities, durations or schedules, so the optimal dose is entirely unknown. Studies should additionally standardise and report stimulation site, laterality, pulse width, frequency, intensity, treatment duration, sham procedures and adherence in accordance with the minimum reporting recommendations proposed by Farmer et al. [43]. Longer blinded follow-up, intention-to-treat analysis and complete adverse-event reporting will be necessary before clinical efficacy can be established.

### 4.8. Strengths

A key strength of this review is the integration of preclinical and clinical evidence, providing a translational overview while retaining separate synthesis and risk-of-bias assessment for each evidence type. Standardised extraction captured study design, stimulation parameters, biological and functional outcomes, safety and treatment adherence. The overlap between the Huguenard and Tan reports was identified and handled explicitly, preventing the NAVSaH cohort from being treated as two independent clinical populations. Quantitative synthesis was restricted to three independent reports providing sufficiently comparable binary functional-outcome data, and the handling of missing outcomes and the post hoc sensitivity analysis are reported transparently. To our knowledge this is the first systematic synthesis focused specifically on haemorrhagic stroke, a population that existing reviews of VNS in stroke have either excluded or represented only as small subgroups.

### 4.9. Limitations

#### 4.9.1. Limitations of the Evidence Base

The evidence base was small, young and heterogeneous. The nine reports represented only four independent clinical cohorts and four preclinical studies, most of which were small pilot or feasibility studies not powered to establish functional efficacy or to detect uncommon adverse events. The total of 96 participants contributing to the meta-analysis is an order of magnitude smaller than would be required to detect a plausible treatment effect on dichotomised mRS, and the resulting estimate is correspondingly imprecise.

The evidence is markedly imbalanced by haemorrhage subtype. Clinical evidence was dominated by SAH, whereas clinical evidence for ICH was limited to the eight-participant haemorrhagic subgroup of Arsava et al. [28], a study designed for safety and feasibility and reporting only 24-hour outcomes. This is a striking translational mismatch, since three of the four preclinical studies used ICH models. The review therefore cannot support any inference about the efficacy of VNS in ICH, and the pooled estimate applies only to aSAH.

Preclinical studies differed substantially in injury model, haemorrhage severity, VNS modality, stimulation protocol and outcome measurement, and, importantly, in comparator: Suzuki et al. used active femoral-nerve stimulation, Cáceres et al. used an inactive device, and Hays et al. and Zhang et al. used rehabilitation alone. These are not equivalent controls, and the last of these cannot separate the effect of stimulation from that of the additional handling and attention received. Methodological reporting was incomplete in several animal studies, particularly for allocation concealment, random housing and blinding, and substantial attrition affected the principal analysis by Hays et al. [25]. All preclinical work used young adult animals of predominantly one sex, limiting relevance to the older, comorbid, mixed-sex populations who sustain most haemorrhagic strokes.

Several limitations attach specifically to the appraisal of the clinical studies. PEDro scores were uniformly high (8–9), which sits uncomfortably alongside the small, exploratory nature of the trials, and reflects limitations of the instrument as much as the quality of the evidence. The subject-blinding item is also poorly calibrated for neurocritical care, where many participants are sedated or have impaired consciousness and are therefore “blinded” trivially rather than by design; conversely, in awake participants, active cervical stimulation produces muscle twitching and lip pulling, as reported by Rebeiz et al. [23], which may unblind participants despite an identical-looking device. Sham adequacy was not formally validated in any included study, and no study reported a blinding-integrity check. A Cochrane risk of bias 2 assessment for the 3 clinical trials of aSAH are included separately in the Supplementary Table 3.

Finally, all included clinical studies reported directionally favourable results. In a field this young, with no published neutral or negative trials, the possibility of publication and outcome-reporting bias cannot be dismissed, even though it could not be assessed formally.

#### 4.9.2. Limitations of the Review Process

The searches were restricted to OVID MEDLINE, while Embase, Web of Science, CENTRAL, CINAHL, grey literature and clinical trial registries were not searched. Restriction to English-language, peer-reviewed publications may have excluded relevant work, particularly from China, where much of the taVNS literature originates.

Several limitations attach to the quantitative synthesis. Favourable outcomes were assessed at different follow-up points across the three contributing reports (discharge for Rebeiz, one month for Myers, first outpatient follow-up for Huguenard), so the pooled estimate combines outcomes measured at different stages of recovery. An I² of 0% should not be read as evidence of true homogeneity, since heterogeneity statistics have very low power with three studies, and REML estimation of τ² is unreliable at this number. Publication bias was not assessed. The severity-based sensitivity analysis was post hoc, altered only the Myers dataset, and did not constitute a formal treatment-by-severity interaction test; it should be regarded as exploratory only.

## 5. Conclusions

This review indicates that VNS is a biologically plausible but currently unproven adjunctive intervention for haemorrhagic stroke. Preclinical studies provided evidence of reduced inflammatory signalling, vascular and structural protection and enhanced neurological recovery, although effects varied according to injury severity and stimulation protocol. Early clinical studies suggest that non-invasive VNS is feasible and reasonably tolerated in patients with SAH, with preliminary signals involving cytokine suppression, autonomic modulation, reduced vasospasm and reduced delayed cerebral ischaemia. The meta-analysis favoured VNS but remained imprecise and non-significant, and the larger effect observed after excluding major haemorrhage was exploratory and requires prospective confirmation. Clinical evidence in ICH remains almost absent despite the strongest preclinical signal arising in ICH models.

The available evidence does not support routine clinical use. It does, however, justify adequately powered multicentre trials that distinguish between acute neuroprotection and rehabilitation-paired treatment, stratify patients by haemorrhage type and severity, standardise and fully report stimulation protocols, prespecify haematoma expansion and infection as outcomes, and assess longer-term, age-appropriate patient-centred outcomes. Given that haemorrhagic stroke is predominantly a disease of later life, such trials should report age-stratified results and record pre-morbid function, frailty and concomitant medication. VNS should therefore be regarded as a promising experimental treatment requiring further clinical validation.

## Supplementary Materials

Table S1: PRISMA 2020 checklist; Table S2: full PubMed search strategy.

## Author Contributions

Conceptualization, A.A.; Methodology, A.A., JB and M.T.; Formal Analysis and data curation, A.A and M.T.; Writing – Original Draft Preparation, A.A. and M.T.; Writing – Review & Editing: A.A, M.T., S.B, M.A., S.L, M.M., F.R., A.M.; Visualization, A.A and M.T.; Supervision, A.A.; All authors have read and agreed to the published version of the manuscript.

## Funding

This research was supported by the NIHR Sheffield Biomedical Research Centre (BRC). The views expressed are those of the authors and not necessarily those of the NHS, the NIHR or the Department of Health and Social Care (DHSC).

## Institutional Review Board Statement

Not applicable. This systematic review analysed previously published data and did not involve new studies of humans or animals.

## Informed Consent Statement

Not applicable.

## Data Availability Statement

The original contributions presented in this study are included in the article and supplementary material. The data extraction spreadsheet and analysis files are available from the corresponding author on reasonable request.

## Conflicts of Interest

Many of the authors of this review, also conducted and authored the Myers et al study and manuscript respectively.

## Notes

### Competing Interest Statement

The authors have declared no competing interest.

### Author Declarations

This systematic review and meta-analysis used data that was freely available from published available databases.

