## Supplementary Table 2 for "Vagus Nerve Stimulation in Intracerebral and Subarachnoid Haemorrhage: A Systematic Review, Narrative Synthesis and Exploratory Meta-Analysis"

### Supplementary Material S1

#### Risk-of-bias assessment of the included randomised controlled trials using the Cochrane risk-of-bias tool for randomised trials (RoB 2)

**Trials assessed:** Huguenard et al. 2025 (NAVSaH); Rebeiz et al. 2024 (VANQUISH); Myers et al. (VNS-SAH).

#### 1. Methods

Risk of bias was assessed with RoB 2, the Cochrane risk-of-bias tool for individually randomised, parallel-group trials[1]. Assessments were made at the level of the individual *result* rather than at the level of the study, because risk of bias in RoB 2 is outcome-specific: within a single trial, different results may sit at different risk of bias. Eighteen results across the three trials were assessed (Table S1.2).

For every result we assessed the effect of *assignment* to intervention (the intention-to-treat effect) rather than the effect of adhering to intervention[2]. Five bias domains were considered: (1) the randomisation process; (2) deviations from intended interventions; (3) missing outcome data; (4) measurement of the outcome; and (5) selection of the reported result. Signalling questions were answered Yes (Y), Probably yes (PY), Probably no (PN), No (N) or No information (NI)[3], and each domain was assigned a judgement of low risk of bias, some concerns, or high risk of bias using the algorithms proposed by the RoB 2 development group. Overall judgements follow the standard rule: low risk only where every domain is low; some concerns where at least one domain raises some concerns and no domain is at high risk; high risk where any domain is at high risk, or where several domains raise some concerns in a way that substantially lowers confidence in the result.

In addition to the primary trial reports, we consulted the published protocol and prospective registry entry for NAVSaH[4]; and the registration identifier reported for VNS-SAH[5]. No registration identifier is reported in the VANQUISH publication and we were unable to identify a corresponding registry record or protocol[6]. Comparison of protocol with report is the principal evidence used in domain 5 and, where a protocol exists, it makes departures from pre-specified analysis detectable. This creates a well-recognised asymmetry in RoB 2: a transparently reported trial with an available protocol can attract a worse domain 5 judgement than a trial whose analysis plan cannot be checked at all. Judgements in this domain should therefore be read alongside the rationale, not as a ranking of research conduct.

**Scope.** RoB 2 addresses internal validity only. It does not capture imprecision arising from small sample sizes, indirectness, publication bias, or funding source and investigator conflicts of interest. All three trials are small pilot or feasibility studies and none was powered for clinical endpoints; these considerations are handled separately under GRADE and are summarised in Section 4. Funding and declared interests are tabulated in Table S1.1 for transparency, not as a bias domain.

---

[1] Sterne JAC, Savović J, Page MJ, et al. RoB 2: a revised tool for assessing risk of bias in randomised trials. BMJ 2019;366:l4898. Assessments follow the full guidance document current at the time of assessment (version of 22 August 2019) and Chapter 8 of the Cochrane Handbook for Systematic Reviews of Interventions, version 6.5.

[2] For harms outcomes the effect of adhering to intervention is sometimes of greater interest. We assessed the effect of assignment throughout so that judgements are directly comparable across trials and outcomes; readers interested in per-protocol effects should note that adherence differed markedly between trials (see Table S1.1).

[3] Y and PY, and N and PN, have the same implications for risk of bias within the RoB 2 algorithms; the distinction records whether the judgement rests on explicit reporting or on inference from the available information. NI is used where the report contains no relevant information and none could be obtained from a protocol or registry entry.

[4] Huguenard AL, Tan G, Johnson G, et al. Non-invasive Auricular Vagus nerve stimulation for Subarachnoid Hemorrhage (NAVSaH): protocol for a prospective, triple-blinded, randomized controlled trial. PLoS One 2024;19(8):e0301154. Registered as NCT04557618 on 21 September 2020; first participant enrolled 4 January 2021, so registration was prospective.

[5] ClinicalTrials.gov NCT06374693; Health Research Authority and Health and Care Research Wales reference 24/YH/0024. The registry record was not independently retrieved at the time of this assessment and the review team should confirm concordance between the registered and reported outcomes before submission.

[6] Absence of a retrievable registry record is not evidence that the trial was unregistered. It does, however, mean that pre-specification of the reported analyses cannot be verified, which is the basis for the domain 5 judgements for that trial.

**Table S1.1. Design features of the included trials relevant to risk of bias**

| Feature | Huguenard 2025 (NAVSaH) | Rebeiz 2024 (VANQUISH) | Myers (VNS-SAH) |
|---|---|---|---|
| **Design and setting** | Prospective, single-centre (Barnes-Jewish Hospital / Washington University, USA), triple-blind, sham-controlled parallel-group RCT; 1:1 allocation. | Multicentre (three Northwell Health hospitals, USA), double-blind, sham-controlled parallel-group RCT; 1:1 allocation. | Prospective, single-centre (Sheffield Teaching Hospitals, UK), sham-controlled parallel-group RCT with blinded outcome assessment; 1:1 allocation stratified by age. |
| **Recruitment period** | January 2021 – October 2023 (64 screened). | October 2019 – June 2022 (120 screened). | April 2024 – September 2025 (73 approached). |
| **Participants randomised** | 27 (taVNS 13; sham 14). Non-traumatic, non-perimesencephalic SAH within 24 h of presentation. | 40 (active 19; sham 21). Spontaneous SAH aged 18–75 y with severe headache (VAS ≥ 7); 4 perimesencephalic. | 30 (taVNS 16; sham 14). Non-traumatic aSAH within 5 days of aneurysm securing. |
| **Intervention** | taVNS to the left concha; 20 min twice daily throughout ICU stay (minimum 7 days); 20 Hz, 250 µs, 0.4 mA (sub-sensory). | Cervical nVNS (gammaCore); two 2-min stimulations up to four times daily, intensity titrated to maximum tolerated (0–40 a.u.); median 9 (IQR 6–12) days. | taVNS to the left tragus (Nurosym, Parasym Ltd); 45 min twice daily for 5 days (10 sessions); 20 Hz, sub-pain-threshold in alert participants and 25 mA in those intubated. |
| **Comparator** | Identical ear clips applied to the left lobule with no current, same duration and schedule. | Visually and acoustically identical inactive device producing no electrical output. | Identical device and settings applied to the left earlobe (mean intensity 23.9 vs 24.4 mA in the active arm). |
| **Blinded** | Participants and families; care providers and medical team; outcome assessors. | Participants; medical team; investigators assessing pain, prescribing analgesia and recording adverse events. | Participants and families; treating clinical teams; research outcome assessors; reporting radiologist. |
| **Not blinded** | Research staff applying the electrodes and setting stimulation parameters (per protocol). | Research assistant who randomised participants, held the allocation log and issued devices. | Research staff delivering the intervention; independent researcher performing randomisation. |
| **Registration / protocol** | NCT04557618, registered prospectively (21 Sep 2020). Full protocol published, including a statistical analysis section. | None reported in the publication; no registry record or protocol identified. IRB approval reported. | NCT06374693; REC 24/YH/0024. No separate protocol or statistical analysis plan publication identified. |
| **Planned vs analysed sample** | 50 planned; enrolment terminated early after an interim analysis on the grounds that the observed effect exceeded that anticipated; 27 analysed. | No sample-size calculation performed ("no statistical hypotheses in this study"); 39 analysed (modified intention-to-treat). | Minimum 12 per group for feasibility; 30 recruited, 29 contributing to outcome tables. |
| **Pre-specified primary endpoints** | Three, per protocol: change in plasma and CSF TNF-α between day 1 and day 13; rate of radiographic vasospasm; and requirement for long-term CSF diversion (ventricular shunt). | Number of serious adverse effects related to nVNS (safety) and compliance (feasibility); main efficacy assessment was change in morphine equivalent dose. | Safety (no taVNS-related SAEs), acceptability (fewer than one third reporting mean side-effect severity ≥ 3) and compliance (> 80% of intended sessions), each with a pre-stated threshold. |
| **Statistical approach** | Linear and generalised linear mixed models; one-sided tests for all outcome variables at α = 0.05; post hoc comparisons at individual time points; no adjustment for multiplicity. | Mixed model repeated measures with no imputation; two-sided tests at α = 0.05; treatment contrasts computed at each of 14 analysis visits; no adjustment for multiplicity. | Mann–Whitney U and chi-square tests on an exploratory, intention-to-treat basis; two-sided α = 0.05; ordinal shift analysis of mRS adjusted for baseline Hunt & Hess grade; no adjustment for multiplicity (acknowledged by the authors). |
| **Funding and declared interests** | Non-commercial (NIH/NINDS, AANS, foundation and institutional grants); funders reported to have had no role. Two authors hold equity in a neuromodulation company and | Funded by the device manufacturer (ElectroCore Inc.), which supplied active and sham devices and supported research staff and journal submission fees; reported not to have been | Non-commercial (NIHR Biomedical Research Centre and University of Sheffield). One author is an employee of the device manufacturer (Parasym), reported not to have been |

| Feature | Huguenard 2025 (NAVSaH) | Rebeiz 2024 (VANQUISH) | Myers (VNS-SAH) |
|---|---|---|---|
| | report pending patents; several report unrelated industry payments. | involved in design, analysis or writing. | involved in study design, data collection or analysis. |
| **Analysis population** | Intention-to-treat for clinical outcomes (27/27); 26/27 for plasma cytokines; 13/27 for CSF cytokines (external ventricular drain required). | Modified intention-to-treat (39/40); one randomised participant who declined all stimulation was excluded. | Intention-to-treat as stated; the participant who withdrew before end of treatment does not appear in the outcome tables (29/30). |

*aSAH, aneurysmal subarachnoid haemorrhage; CSF, cerebrospinal fluid; ICU, intensive care unit; IQR, interquartile range; mRS, modified Rankin Scale; nVNS, non-invasive (cervical) vagus nerve stimulation; RCT, randomised controlled trial; SAE, serious adverse event; SAH, subarachnoid haemorrhage; taVNS, transauricular vagus nerve stimulation; TNF-α, tumour necrosis factor alpha; VAS, visual analogue scale.*

#### 2. Summary of risk-of-bias judgements

Table S1.2 records the domain-level and overall judgement for each assessed result. Domains are: D1 randomisation process; D2 deviations from intended interventions; D3 missing outcome data; D4 measurement of the outcome; D5 selection of the reported result. Detailed signalling-question responses and rationales are given in Section 3[7].

**Table S1.2. RoB 2 judgements for each assessed result**

| Trial and result assessed | D1 | D2 | D3 | D4 | D5 | Overall |
|---|---|---|---|---|---|---|
| **Huguenard et al. 2025 — NAVSaH (n = 27)** | | | | | | |
| R1. Moderate or severe radiographic vasospasm (blinded clinician assessment, participant level) | Low | Low | Low | Some concerns | High | High |
| R2. Quantitative vasospasm: vessel-level severity and normalised vessel calibre over time | Low | Low | Some concerns | Some concerns | High | High |
| R3. Plasma TNF-α and IL-6 | Low | Low | Some concerns | Low | High | High |
| R4. CSF TNF-α and IL-6 | Low | Low | High | Low | High | High |
| R5. Functional outcome: mRS at discharge and first follow-up; discharge destination | Low | Low | Low | Some concerns | High | High |
| R6. Adverse events | Low | Low | Low | Some concerns | Some concerns | Some concerns |
| **Rebeiz et al. 2024 — VANQUISH (n = 40)** | | | | | | |
| R7. Serious adverse events (primary safety endpoint) | Low | Some concerns | Low | Low | Some concerns | Some concerns |
| R8. Non-serious device-related side effects | Low | Some concerns | Low | Some concerns | Some concerns | Some concerns |
| R9. Headache intensity (VAS, pre- to post-stimulation) | Low | Some concerns | Some concerns | High | High | High |
| R10. Morphine equivalent dose at days 7 and 14 | Low | Some concerns | Some concerns | Some concerns | Some concerns | Some concerns |
| R11. New cerebral infarction on neuroimaging | Low | Some concerns | Low | Some concerns | Some concerns | Some concerns |

7 The judgements in this table can be exported directly into a traffic-light figure for the main report using robvis, which accepts the domain codes used here. Where an overall judgement of high risk arises from a single domain, that domain is identified in the rationale so that readers can see what drives it.

| Trial and result assessed | D1 | D2 | D3 | D4 | D5 | Overall |
|---|---|---|---|---|---|---|
| R12. mRS 0–2 at hospital discharge | Low | Some concerns | Some concerns | Some concerns | Some concerns | Some concerns |
| **Myers et al. — VNS-SAH (n = 30)** | | | | | | |
| R13. Safety: taVNS-related serious adverse events | Low | Low | Low | Low | Low | Low |
| R14. Compliance: proportion of intended sessions delivered | Low | Low | Low | Low | Low | Low |
| R15. Acceptability: participant-rated side-effect severity | Low | Low | Some concerns | Low | Low | Some concerns |
| R16. Change in serum inflammatory mediators (TNF-α, IL-1, IL-6, IL-10, CRP) | Low | Low | Some concerns | Low | Some concerns | Some concerns |
| R17. Delayed cerebral ischaemia | Low | Low | Low | Some concerns | Some concerns | Some concerns |
| R18. mRS at 1 month (ordinal shift analysis) | Low | Low | Low | Low | Some concerns | Some concerns |

*CRP, C-reactive protein; CSF, cerebrospinal fluid; IL, interleukin; mRS, modified Rankin Scale; TNF-α, tumour necrosis factor alpha; VAS, visual analogue scale. Green = low risk of bias; amber = some concerns; red = high risk of bias.*

#### 3. Detailed domain-by-domain assessments

##### 3.1 Huguenard et al. 2025 — NAVSaH

*Auricular vagus nerve stimulation for mitigation of inflammation and vasospasm in subarachnoid hemorrhage: a single-institution randomized controlled trial.* J Neurosurg 2025;142:1720–31. Twenty-seven participants with spontaneous SAH randomised to transauricular vagus nerve stimulation (n = 13) or sham stimulation (n = 14)8.

**Table S1.3. RoB 2 signalling questions — NAVSaH**

| Signalling question | Response | Supporting evidence and rationale |
|---|---|---|
| **Domain 1. Bias arising from the randomisation process** | | |
| 1.1 Was the allocation sequence random? | Y | Simple randomisation using computer-generated random numbers, described consistently in the trial report and the published protocol. |
| 1.2 Was the allocation sequence concealed until participants were enrolled and assigned to interventions? | Y | The next allocation was concealed until consent and enrolment were complete; a single investigator entered the participant into the system to obtain the assignment, and eligible patients were allocated strictly in sequence. |
| 1.3 Did baseline differences between intervention groups suggest a problem with the randomisation process? | PN | No significant baseline differences in sex, race, aneurysm treatment modality, age, Hunt & Hess grade, modified Fisher score, Glasgow Coma Scale score or admission mRS. Numerical differences (mean age 62.9 vs 56.9 years; male sex 30.8% vs 14.3%) are of the magnitude expected by chance with 27 participants and do not indicate a failure of the randomisation process. |
| **Domain judgement** | Low | *Applies to all six results. Sequence generation and concealment are adequately described and baseline data are compatible with successful randomisation.* |
| **Domain 2. Bias due to deviations from intended interventions (effect of assignment to intervention)** | | |
| 2.1 Were participants aware of their assigned intervention during the trial? | PN | Stimulation parameters were sub-sensory (0.4 mA) and both arms received identical ear clips for identical durations and at identical times. |

8 Assessment was made on the final journal version. An earlier preprint of the same analysis is available (medRxiv 10.1101/2024.04.29.24306598), and further outcomes from the same 27 participants have been reported separately — continuous cardiovascular physiology (eLife 2025;13:RP100088) and a hospital cost analysis. Reviewers should ensure that results from these companion publications are attributed to the single NAVSaH randomised comparison rather than counted as separate studies.

| Signalling question | Response | Supporting evidence and rationale |
|---|---|---|
| | | The authors note that electrode position differed between arms (concha vs lobule) and that this could in principle allow a group difference to be detected, but not which position was active. |
| 2.2 Were carers and people delivering the interventions aware of participants' assigned intervention? | **Y (intervention providers) / PN (clinical team)** | Per protocol, research staff applying the clips and setting stimulation parameters were not blinded. The treating medical team, who made all management decisions, and the outcome assessors were blinded. The authors acknowledge that clinician presumption of allocation could in principle have influenced other aspects of care. |
| 2.3 Were there deviations from the intended intervention that arose because of the trial context? | **PN** | No protocol deviations are reported. One participant in each arm discontinued treatment sessions and sampling before completion. Interventions for vasospasm (blood-pressure augmentation, intra-arterial or intrathecal vasodilators, angioplasty) were numerically less frequent in the taVNS arm, but these are responses to the outcome rather than trial-driven deviations, and none of the between-group differences reached significance. |
| 2.4 / 2.5 Were these deviations likely to have affected the outcome, and were they balanced between groups? | **Not applicable** | Not reached, given the response to 2.3. |
| 2.6 Was an appropriate analysis used to estimate the effect of assignment to intervention? | **Y (clinical outcomes) / PN (plasma cytokines)** | Clinical outcomes were analysed by intention to treat in all 27 randomised participants. One randomised participant was excluded from the cytokine analyses because only a single baseline sample had been obtained. |
| 2.7 Was there potential for a substantial impact of the failure to analyse participants in the group to which they were randomised? | **PN** | The exclusion concerns one of 27 participants for whom no post-baseline outcome data existed; the issue is more properly considered under domain 3. |
| **Domain judgement** | **Low** | *Applies to all six results. Blinding of participants and of the clinical team was plausibly maintained, no trial-context deviations are identified, and analysis was by assigned group.* |
| **Domain 3. Bias due to missing outcome data** | | |
| 3.1 Were data for this outcome available for all, or nearly all, participants randomised? | **Y (R1, R5, R6) / PY (mRS at follow-up) / PN (R2, R3) / N (R4)** | Vasospasm, discharge destination and adverse events: 27/27. mRS at first follow-up: 25/27 (one participant lost to follow-up in each arm). Plasma cytokines: 26/27 at baseline, with progressive loss of later time points as participants left the intensive care unit; the number contributing at each time point is not reported. CSF cytokines: 13/27 (6 taVNS, 7 sham), because sampling required an external ventricular drain to be in situ. |
| 3.2 Is there evidence that the result was not biased by missing outcome data? | **N (R2–R4)** | No sensitivity analysis, imputation, or comparison of participants with and without complete data is reported. The mixed models used assume that data are missing at random. |
| 3.3 Could missingness in the outcome depend on its true value? | **PY (R2–R4)** | Availability of CSF depends on the presence of an external ventricular drain, which is determined by hydrocephalus and haemorrhage severity — factors strongly associated with cytokine concentrations and with vasospasm. Availability of later blood samples and later imaging depends on length of intensive care stay, which is likewise severity-dependent. |
| 3.4 Is it likely that missingness in the outcome depended on its true value? | **PY (R4) / PN (R2, R3)** | The CSF findings rest on comparisons at day 13, which can only include participants whose drain remained in situ at that point; the numbers contributing are not reported and are necessarily a subset of the 13 with any CSF sampling. For plasma, more than 95% of participants contributed a baseline and at least one on-treatment sample, so a comparable dependence is less likely. |
| **Domain judgement** | **See rationale** | **Low risk** for R1, R5 and R6. **Some concerns** for R2 and R3 (progressive, unreported attrition across time points with no sensitivity analysis). **High risk** for R4: fewer than half of randomised participants could contribute CSF at all, and eligibility for CSF sampling is determined by the same disease severity that drives the outcome. |
| **Domain 4. Bias in measurement of the outcome** | | |

| Signalling question | Response | Supporting evidence and rationale |
|---|---|---|
| 4.1 Was the method of measuring the outcome inappropriate? | **N (R1–R4) / PN (R5, R6)** | Cytokines were measured by immunoassay on samples processed immediately. Vasospasm was graded using conventional radiological thresholds (< 25%, 25–50%, > 50% narrowing) with calibrated serial vessel measurements normalised to the baseline study. Discharge destination is a legitimate outcome in its own right but is a weak proxy for neurological function, being influenced by rehabilitation bed availability, insurance status and family circumstances. |
| 4.2 Could measurement or ascertainment of the outcome have differed between intervention groups? | **NI (R1, R2, R5, R6) / PN (R3, R4)** | All participants underwent a protocolised vascular study at approximately day 7, but further imaging was performed at the discretion of the clinical teams when vasospasm or stroke was suspected. The mean number of vascular studies differed (2.8 [SD 0.8] taVNS vs 3.4 [SD 1.2] sham; p = 0.28). A participant-level endpoint defined as vasospasm on any imaging study is sensitive to how many studies are obtained, so unequal ascertainment intensity could influence the comparison. Because the ordering clinicians were blinded, this difference may equally be a consequence of the intervention rather than a source of bias; the report does not allow the two explanations to be separated. Adverse-event ascertainment is not described (no schedule, instrument or definitions are given). |
| 4.3 Were outcome assessors aware of the intervention received? | **PN** | Imaging was reported by a neuroradiologist or endovascular neurointerventionalist blinded to allocation, and all quantitative measurements were made by a single blinded, fellowship-trained reviewer who was also blinded to participant identity and to the indication for the study. mRS scores were assigned by blinded assessors. |
| 4.4 / 4.5 Could and was assessment of the outcome influenced by knowledge of intervention received? | **PN** | Not reached in substance, given the response to 4.3. |
| **Domain judgement** | **See rationale** | **Low risk** for R3 and R4 (objective laboratory measurement, blinded processing). **Some concerns** for R1, R2, R5 and R6, reflecting unequal imaging intensity between arms for an "any study" endpoint, reliance on discharge destination as a functional proxy determined by the treating service, and the absence of any description of adverse-event ascertainment. |
| **Domain 5. Bias in selection of the reported result** | | |
| 5.1 Were the data producing this result analysed in accordance with a pre-specified analysis plan finalised before unblinded outcome data were available? | **PN** | A protocol containing a statistical analysis section was published. The reported analyses depart from it in three respects. (i) The protocol specified that the TNF-α effect would be examined through the time-by-treatment interaction from day 1 to day 13; the report instead presents one-sided post hoc comparisons at individual time points (days 4, 7, 10 and 13) and does not report the pre-specified interaction test. (ii) The protocol specified a mixed-effects logistic regression with generalised estimating equations for the binary vasospasm endpoint; the report presents chi-square and analysis-of-variance comparisons. (iii) All outcome analyses are reported as one-sided, whereas the protocol's power calculations specified two-sided tests at α = 0.05. |
| 5.2 Is the numerical result likely to have been selected, on the basis of the results, from multiple eligible outcome measurements within the outcome domain? | **PY (R1–R5) / PN (R6)** | A panel of 13 cytokines was assayed per protocol; two analytes are reported, at five time points, with emphasis on the time points reaching significance. Vasospasm is reported in four different ways (any vasospasm; moderate or severe by clinician assessment; vessel-level severity categories; normalised calibre over time). Functional outcome is reported as mean mRS, as dichotomised good outcome, as change from admission, and as discharge destination. The third pre-specified primary endpoint — requirement for long-term CSF diversion — is not reported at all. All adverse-event data are reported (none occurred), so 5.2 does not apply to R6. |
| 5.3 Is the numerical result likely to have been selected, on the basis of the results, from multiple eligible analyses of the data? | **PY (R1–R5) / PN (R6)** | Participant-level and vessel-level analyses of the same construct are both presented; the vessel-level comparison treats 164 and 188 vessels as independent units. For functional outcome, within-group pre-post comparisons (taVNS p = 0.014; sham p = 0.18) are emphasised after the pre-specified between-group interaction was non-significant (p = 0.20) — a comparison of significance rather than a test of the difference between groups. |

| Signalling question | Response | Supporting evidence and rationale |
|---|---|---|
| **Domain judgement** | **See rationale** | **High risk** for R1–R5. The reported analyses depart from the pre-specified plan in ways that increase the chance of a positive finding, one of three pre-specified primary endpoints is unreported, and the outcome constructs are each presented in multiple forms. **Some concerns** for R6, where the departure from the pre-specified plan applies but complete reporting of adverse events removes the scope for selection. |

***Additional considerations recorded during assessment.*** *(a) Enrolment was stopped early, after an interim analysis, on the grounds that the observed reduction in moderate or severe vasospasm (> 40%) exceeded the 30% anticipated; no pre-specified stopping boundary or alpha-spending function is described, and trials stopped early for benefit systematically overestimate treatment effects. (b) Because all outcome p values are one-sided, the corresponding two-sided values are approximately double; on that basis several results reported as significant would not meet a conventional two-sided threshold of 0.05, including any radiographic vasospasm (p = 0.035), plasma TNF-α on day 10 (p = 0.030), CSF TNF-α on day 13 (p = 0.031) and poor discharge destination (p = 0.04). Exact two-sided values are not reported. (c) The vessel-level analysis compares 164 vessels in 13 participants with 188 vessels in 14 participants using a chi-square test; treating vessels as independent overstates precision unless clustering within participants is modelled. (d) Eligibility criteria differ between protocol and report (for example, the bradycardia exclusion is a heart rate below 50 beats per minute for more than 5 minutes in the protocol and below 40 beats per minute for more than 10 minutes in the report).*

#### 3.2 Rebeiz et al. 2024 — VANQUISH

*Noninvasive vagus nerve stimulation in spontaneous subarachnoid hemorrhage (VANQUISH): a randomized safety and feasibility study.* Brain Stimul 2024;17:543–9. Forty participants with spontaneous SAH randomised to cervical non-invasive vagus nerve stimulation (n = 19) or sham (n = 21); 39 analysed9.

**Table S1.4. RoB 2 signalling questions — VANQUISH**

| Signalling question | Response | Supporting evidence and rationale |
|---|---|---|
| **Domain 1. Bias arising from the randomisation process** | | |
| 1.1 Was the allocation sequence random? | **Y** | Randomisation used a variable block design with 1:1 allocation. |
| 1.2 Was the allocation sequence concealed until participants were enrolled and assigned to interventions? | **PY** | Allocation concealment is stated. Active and sham devices were identical in design, shape, colour and operation and were distinguishable only by serial number; an unblinded research assistant held the randomisation log and, once device training was complete, had no further contact with participants or clinicians. |
| 1.3 Did baseline differences between intervention groups suggest a problem with the randomisation process? | **PN** | Baseline characteristics were similar for age, sex, Hunt & Hess classification, modified Fisher classification, treatment modality and SAH aetiology. |
| **Domain judgement** | **Low** | *Applies to all six results.* |
| **Domain 2. Bias due to deviations from intended interventions (effect of assignment to intervention)** | | |
| 2.1 Were participants aware of their assigned intervention during the trial? | **PY** | Although the sham device was visually and acoustically identical, active stimulation intensity was titrated upward to the maximum tolerated level and produced perceptible effects significantly more often than sham: muscle twitching 44.4% vs 9.5% (p = 0.02), lip pull 33.3% vs 0% (p = 0.005) and nausea or vomiting 22.2% vs 0% (p = 0.03). Participants in the active arm could therefore frequently infer their allocation, and the authors acknowledge that this may have introduced bias. |
| 2.2 Were carers and people delivering the interventions aware of participants' assigned intervention? | **PY** | Nursing and clinical staff administered, supervised and assisted with each session at the bedside and would have observed the same visible effects; the same staff recorded pain scores and administered analgesia. |
| 2.3 Were there deviations from the intended intervention that arose because of the trial context? | **NI** | Of 1087 attempted stimulations, 172 (19%) were declined by participants, in part because they felt the device was not helping; a further 133 of 1260 attempts (12%) were withheld for sinus bradycardia and 40 (3%) were not delivered for logistical reasons. |

9 The trial was designed as a safety and feasibility study; the authors state explicitly that there were no statistical hypotheses and that no sample-size calculation was performed. All efficacy comparisons are therefore exploratory by design, and the risk-of-bias judgements for those results should be read in that light rather than as an assessment of a failed efficacy trial.

| Signalling question | Response | Supporting evidence and rationale |
|---|---|---|
| | | Adherence is not reported separately by arm, so it cannot be determined whether trial-context deviations differed between groups. |
| 2.4 Were these deviations likely to have affected the outcome? | **PY** | Analgesia was nurse-administered according to a unit protocol but titrated to reported pain. Awareness of allocation could plausibly influence both requests for and administration of opioids, which is the trial's main efficacy outcome, as well as reported pain itself. |
| 2.5 Were these deviations balanced between groups? | **NI** | Per-arm figures are not reported. |
| 2.6 Was an appropriate analysis used to estimate the effect of assignment to intervention? | **PN** | The primary analysis used a modified intention-to-treat population; one randomised participant who declined all stimulation after consent was excluded. |
| 2.7 Was there potential for a substantial impact of the failure to analyse participants in the group to which they were randomised? | **PN** | The exclusion concerns one of 40 participants and is unlikely, by itself, to have materially changed the estimates. |
| **Domain judgement** | **Some concerns** | *Applies to all six results. Blinding of participants and bedside staff was substantially compromised by the perceptible side-effect profile of the active device, adherence is not reported by arm, and one randomised participant was excluded from analysis.* |
| **Domain 3. Bias due to missing outcome data** | | |
| 3.1 Were data for this outcome available for all, or nearly all, participants randomised? | **PY (R7, R8, R11) / PN (R9, R10, R12)** | Safety endpoints and cerebral infarction are reported for the full modified intention-to-treat population (18 and 21). The pain and morphine-equivalent-dose analyses extend to day 14, whereas the median duration of stimulation was 9 days (IQR 6–12), so the number contributing falls steeply across the analysed period. For mRS at discharge the denominators are inconsistent between the flow diagram and the text. |
| 3.2 Is there evidence that the result was not biased by missing outcome data? | **N** | No imputation was used and no sensitivity analysis or comparison of completers with non-completers is reported; the mixed model repeated measures analysis assumes data are missing at random. |
| 3.3 Could missingness in the outcome depend on its true value? | **PY** | Discontinuation of stimulation and discharge from hospital are driven by clinical course and by pain itself, so later observations are missing in a manner related to the true values of the outcomes. |
| 3.4 Is it likely that missingness in the outcome depended on its true value? | **PN** | The principal pain finding is a within-session pre-to-post contrast among sessions actually delivered, which is less exposed to later attrition than the day 7 and day 14 dose comparisons. |
| **Domain judgement** | **See rationale** | **Low risk** for R7, R8 and R11. **Some concerns** for R9, R10 and R12, reflecting steep and unquantified attrition across the analysed time course and inconsistent denominators for the discharge mRS. |
| **Domain 4. Bias in measurement of the outcome** | | |
| 4.1 Was the method of measuring the outcome inappropriate? | **N** | Visual analogue scale pain scores, morphine equivalent dose derived by standard conversion factors, mRS and imaging-defined infarction are all appropriate measures of their respective constructs. |
| 4.2 Could measurement or ascertainment of the outcome have differed between intervention groups? | **PN (R7, R9, R10, R12) / PY (R8) / NI (R11)** | Serious adverse events were captured by continuous telemetry and standard clinical monitoring applied identically in both arms. Non-serious device-related effects are by design expected to differ, since they are direct consequences of active stimulation; this is a true effect rather than a bias, but it means the measure cannot be interpreted independently of unblinding. Cerebral infarction was ascertained by comparing the last available neuroimaging study with the first study obtained after aneurysm treatment; imaging was not protocolised, so the number and timing of scans could differ between arms. |
| 4.3 Were outcome assessors aware of the intervention received? | **Y (R9) / NI (R11, R12) / PN (R7, R10)** | For pain the outcome assessor is the participant, who could frequently infer allocation from perceptible stimulation effects. Blinded adjudication is not described for cerebral infarction or for mRS at discharge; the report states only that investigators and clinicians who assessed pain intensity, provided analgesia and monitored adverse events were blinded. |

| Signalling question | Response | Supporting evidence and rationale |
|---|---|---|
| 4.4 Could assessment of the outcome have been influenced by knowledge of intervention received? | **Y (R9) / PN (R11, R12)** | A self-reported subjective score recorded immediately before and after a perceptible intervention is highly susceptible to expectation effects. Imaging-defined infarction and mRS are less susceptible, although not immune. |
| 4.5 Is it likely that assessment of the outcome was influenced by knowledge of intervention received? | **PY (R9) / PN (others)** | The trial's positive efficacy signal is a within-session pre-to-post change in a subjective score — precisely the measurement most vulnerable to unblinding — while the more objective co-primary efficacy measure, morphine equivalent dose, showed no between-group difference at day 7 (p = 0.93), day 14 (p = 0.79) or overall (p = 0.66). This pattern is consistent with, though not proof of, a measurement effect. |
| **Domain judgement** | **See rationale** | **Low risk** for R7. **Some concerns** for R8, R10, R11 and R12. **High risk** for R9: a subjective, self-reported outcome assessed by participants whose blinding was demonstrably incomplete. |
| **Domain 5. Bias in selection of the reported result** | | |
| 5.1 Were the data producing this result analysed in accordance with a pre-specified analysis plan finalised before unblinded outcome data were available? | **NI** | No trial registration identifier is reported in the publication and no registry record, protocol or statistical analysis plan could be identified. It is therefore not possible to verify whether the reported outcomes and analyses were pre-specified. The report states that there were no statistical hypotheses and that no sample-size calculation was performed. |
| 5.2 Is the numerical result likely to have been selected, on the basis of the results, from multiple eligible outcome measurements within the outcome domain? | **PY (R9) / PN (R7, R8, R10, R11, R12)** | Pain is reported in several eligible forms — mean post-stimulation intensity, the pre-to-post difference, within-group and between-group comparisons, and daily values across 15 days — and the headline efficacy claim rests on one of these. By contrast, all eleven safety endpoints and all pre-stated exploratory endpoints are reported irrespective of direction, including null and unfavourable results, which reduces the scope for selective reporting of those results. |
| 5.3 Is the numerical result likely to have been selected, on the basis of the results, from multiple eligible analyses of the data? | **PY (R9) / PN (others)** | Treatment-group contrasts were computed at each of 14 analysis visits in addition to the pooled comparisons, without adjustment for multiplicity. |
| **Domain judgement** | **See rationale** | **Some concerns** for R7, R8, R10, R11 and R12: pre-specification cannot be verified, but reporting within each outcome domain appears complete. **High risk** for R9, where an unverifiable analysis plan is combined with multiple eligible measurements and analyses of a subjective outcome. |

***Additional considerations recorded during assessment.*** *(a) The trial was funded by the manufacturer of the investigational device, which supplied both active and sham devices and met journal submission fees; the report states the company had no role in design, analysis or writing. RoB 2 contains no funding domain, but this should be reported alongside the risk-of-bias table. (b) Percentages for neurological deterioration attributed to vasospasm (3 [19%] active vs 6 [31%] sham) are not reconcilable with the stated analysis denominators of 18 and 21, and the denominator for mRS at discharge in the sham arm is given as 20 in the text and 21 in the flow diagram; data extraction for the review should record the counts rather than the percentages. (c) Because the active device produces perceptible muscle twitching and lip pull, blinding is difficult to achieve with an inert sham; future trials of cervical nVNS may require an active sham delivering sub-threshold stimulation, or blinding assessment at the end of treatment.*

#### 3.3 Myers et al. — VNS-SAH

*Transauricular vagus nerve stimulation for aneurysmal subarachnoid haemorrhage (VNS-SAH): a pilot randomised controlled trial.* Thirty participants with aneurysmal SAH randomised to transauricular vagus nerve stimulation (n = 16) or sham stimulation (n = 14) within 5 days of aneurysm securing[10].

**Table S1.5. RoB 2 signalling questions — VNS-SAH**

| Signalling question | Response | Supporting evidence and rationale |
|---|---|---|
| **Domain 1. Bias arising from the randomisation process** | | |

10 Assessed on the manuscript draft dated 30 August 2026. Judgements should be re-checked against the published version, since table numbering, denominators and any additional analyses requested at peer review may change.

| Signalling question | Response | Supporting evidence and rationale |
|---|---|---|
| 1.1 Was the allocation sequence random? | **Y** | Block randomisation with 1:1 allocation, stratified by age (< 65 and ≥ 65 years), generated by a validated external online randomisation service. |
| 1.2 Was the allocation sequence concealed until participants were enrolled and assigned to interventions? | **Y** | Allocation was performed by an independent researcher using a central web-based system after enrolment, so the upcoming assignment could not be foreseen by the recruiting team. |
| 1.3 Did baseline differences between intervention groups suggest a problem with the randomisation process? | **PN** | The taVNS arm had numerically more severe haemorrhage (Hunt & Hess grade 5 in 25% vs 0%; grade 1 in 0% vs 21.4%; distribution p = 0.082), more participants intubated and ventilated (37.5% vs 28.6%) and a higher white cell count. With central concealed allocation and 30 participants this degree of imbalance is compatible with chance and does not indicate a failure of the randomisation process. It is nonetheless prognostically important and should be handled as confounding at the interpretation stage, not as bias in this domain; the imbalance runs against the intervention arm. |
| **Domain judgement** | **Low** | *Applies to all six results.* |
| **Domain 2. Bias due to deviations from intended interventions (effect of assignment to intervention)** | | |
| 2.1 Were participants aware of their assigned intervention during the trial? | **PN** | Both arms received current from the same device at closely similar intensities (mean 24.4 mA [SD 4.8] active vs 23.9 mA [SD 4.3] sham), differing only in electrode site (tragus vs earlobe). The reported side-effect profile does not suggest that participants could identify allocation: headache was reported more often in the sham arm (90% vs 40%) and nausea occurred only in the sham arm. |
| 2.2 Were carers and people delivering the interventions aware of participants' assigned intervention? | **Y (intervention providers) / PN (clinical team and assessors)** | Research staff delivering the intervention were unblinded by design. Participants and families, the treating clinical teams who made all management decisions, the research outcome assessors and the reporting radiologist were blinded. The intervention providers had no role in clinical management or outcome assessment. |
| 2.3 Were there deviations from the intended intervention that arose because of the trial context? | **PN** | No protocol deviations are reported. All participants received nimodipine as standard care. One participant in the taVNS arm withdrew after three sessions because of headache; the pre-treatment headache score in that participant was already high (7/10). Session delivery was high and similar overall (221 of 259 possible sessions, 85.3%, of which 215 were of full duration). |
| 2.4 / 2.5 Were these deviations likely to have affected the outcome, and were they balanced between groups? | **Not applicable** | Not reached, given the response to 2.3. |
| 2.6 Was an appropriate analysis used to estimate the effect of assignment to intervention? | **PY** | Analyses are described as intention to treat. The participant who withdrew consent before end of treatment is not represented in the outcome tables (taVNS n = 15 rather than 16), but outcome data for that participant were not available rather than being available and excluded; this is treated under domain 3. |
| 2.7 Was there potential for a substantial impact of the failure to analyse participants in the group to which they were randomised? | **PN** | One participant of 30; for the count outcomes, inclusion or exclusion changes proportions by a few percentage points and would not alter the direction of any comparison. |
| **Domain judgement** | **Low** | *Applies to all six results. The sham delivers matched stimulation at a non-vagal site, blinding of participants and clinicians appears to have been maintained, and unblinded staff had no role in management or assessment.* |
| **Domain 3. Bias due to missing outcome data** | | |
| 3.1 Were data for this outcome available for all, or nearly all, participants randomised? | **Y (R13, R14) / PY (R17, R18) / N (R15, R16)** | Safety and compliance data cover all randomised participants and all possible sessions. One-month follow-up data were available for 29 of 30 (96.7%). Paired baseline and end-of-treatment inflammatory marker samples were available for 23 of 30 (76.7%). Acceptability ratings could only be obtained from participants alert enough to provide them, whereas 10 of 30 (33.3%) were intubated and ventilated at randomisation. |

| Signalling question | Response | Supporting evidence and rationale |
|---|---|---|
| 3.2 Is there evidence that the result was not biased by missing outcome data? | **N (R15, R16)** | No sensitivity analysis, imputation or comparison of participants with and without paired samples is reported. |
| 3.3 Could missingness in the outcome depend on its true value? | **PY (R15, R16)** | Absent end-of-treatment samples arise principally from early inpatient death, early discharge and withdrawal, all of which relate to haemorrhage severity and therefore to systemic inflammatory marker concentrations. Three of the four inpatient deaths occurred in the taVNS arm (mean Hunt & Hess grade 4.7), so the sample loss may not be balanced with respect to severity. Ability to report side-effect severity depends on conscious level, which is itself related to tolerability and to how stimulation is experienced. |
| 3.4 Is it likely that missingness in the outcome depended on its true value? | **PN** | Roughly three quarters of randomised participants contributed paired samples, and the direction of any resulting bias is not predictable from the available data; the losses plausibly attenuate rather than create the observed between-group difference, since the arm with more early deaths is the arm showing greater reduction in TNF-α. |
| **Domain judgement** | **See rationale** | **Low risk** for R13, R14, R17 and R18. **Some concerns** for R15 and R16. For acceptability, the judgement depends on how the outcome is defined: if acceptability is a property of all randomised participants, roughly a third could not contribute and the domain would be at high risk; if it is defined among participants able to self-report, the data are effectively complete and the limitation is one of applicability rather than bias. We have taken the second reading, consistent with how the outcome is operationalised in the report, and flag the restriction as an indirectness issue for GRADE. |
| **Domain 4. Bias in measurement of the outcome** | | |
| 4.1 Was the method of measuring the outcome inappropriate? | **N / PN** | Delayed cerebral ischaemia was defined using the SAHIT consensus definition. mRS and Barthel Index are standard instruments. Serum mediators were assayed on samples processed immediately. Acceptability used a pre-specified 5-point Likert rating of named expected side effects after each session, with a pre-stated threshold. |
| 4.2 Could measurement or ascertainment of the outcome have differed between intervention groups? | **PN (R13–R16, R18) / NI (R17)** | Clinical detection of delayed cerebral ischaemia requires observation of a new focal deficit or a fall in Glasgow Coma Score of at least 2 points. The taVNS arm contained more participants who were intubated and ventilated (37.5% vs 28.6%) and experienced more early inpatient deaths (3 vs 1), both of which reduce the opportunity to observe and record clinical deterioration, and death acts as a competing event for the observation of delayed cerebral ischaemia. The imaging schedule is not stated to have been protocolised, although all neuroimaging was reviewed by a radiologist blinded to allocation. The reported difference (6.6% vs 35.7%) is therefore susceptible to differential ascertainment in a direction that is not predictable a priori. |
| 4.3 Were outcome assessors aware of the intervention received? | **PN** | Outcome measures were collected by researchers blinded to allocation and to intervention delivery, and neuroimaging was reported by a blinded radiologist. Adverse events were recorded by the responsible clinical teams, who were blinded. Acceptability was rated by participants, who were blinded. |
| 4.4 / 4.5 Could and was assessment of the outcome influenced by knowledge of intervention received? | **PN** | Not reached in substance, given the response to 4.3. |
| **Domain judgement** | **See rationale** | **Low risk** for R13–R16 and R18. **Some concerns** for R17, reflecting unequal opportunity to ascertain a clinically defined outcome in an arm with more sedated participants and more early deaths, and the absence of a stated imaging schedule. A competing-risk-aware analysis, or reporting of delayed cerebral ischaemia among participants surviving and assessable to day 14, would address this. |
| **Domain 5. Bias in selection of the reported result** | | |
| 5.1 Were the data producing this result analysed in accordance with a pre-specified analysis plan finalised before unblinded outcome data were available? | **PY (R13–R15) / NI (R16–R18)** | The trial was prospectively registered and the three primary feasibility outcomes are reported against explicit, quantitative thresholds stated in advance (no taVNS-related serious adverse events; fewer than one third of participants reporting mean side-effect severity ≥ 3; more than |

| Signalling question | Response | Supporting evidence and rationale |
|---|---|---|
| | | 80% of intended sessions delivered). For the secondary outcomes, the report describes the analysis methods used but does not state that a statistical analysis plan was finalised before unblinding, and the ordinal shift analysis adjusted for baseline Hunt & Hess grade is not explicitly identified as pre-specified. |
| 5.2 Is the numerical result likely to have been selected, on the basis of the results, from multiple eligible outcome measurements within the outcome domain? | **PN** | All five measured inflammatory mediators are reported with their p values irrespective of direction, including null results (IL-6 p = 0.798; CRP p = 0.762) and results in the unexpected direction (IL-10). All pre-stated secondary clinical outcomes appear in Table 2, including those favouring sham. Functional outcome is reported as median mRS, as a dichotomy and as an ordinal shift. |
| 5.3 Is the numerical result likely to have been selected, on the basis of the results, from multiple eligible analyses of the data? | **PN** | A single analytic approach is applied consistently across outcomes. No adjustment for multiplicity was made across the panel of five mediators and the range of secondary outcomes, which the authors explicitly acknowledge as a limitation; with roughly twenty secondary comparisons, one or two nominally significant results would be expected by chance alone, and the TNF-α result (p = 0.028) should be interpreted accordingly. |
| **Domain judgement** | **See rationale** | **Low risk** for R13, R14 and R15 (pre-registered primary outcomes with pre-stated thresholds). **Some concerns** for R16, R17 and R18: reporting is complete and even-handed, but pre-specification of the secondary analyses cannot be verified and no allowance is made for multiplicity. |

***Additional considerations recorded during assessment.*** *(a) The trial is described as single-blind, but the blinding actually described — of participants, families, treating teams, outcome assessors and the reporting radiologist, with only the staff delivering the intervention unblinded — corresponds to what most trials would report as double or triple blinding, and is the same configuration as the trial reported as triple-blind by Huguenard and colleagues. The label understates the design. (b) The confidence interval around the one-month ordinal shift estimate (common odds ratio 0.83, 95% CI 0.16–4.24) spans effects from substantial benefit to substantial harm; this is imprecision rather than bias and belongs under GRADE. (c) Consider pre-specifying and reporting delayed cerebral ischaemia with an explicit accounting for death as a competing event in any subsequent trial.*
