## Supplementary Table 3 for "Vagus Nerve Stimulation in Intracerebral and Subarachnoid Haemorrhage: A Systematic Review, Narrative Synthesis and Exploratory Meta-Analysis"

### Supplementary File 2. Search strategies for Ovid MEDLINE

Searches were built around two concepts, combined with the Boolean operator AND: (1) haemorrhagic stroke, intracerebral haemorrhage, subarachnoid haemorrhage and intracranial aneurysm; and (2) vagus nerve stimulation. Each concept combined controlled vocabulary headings with free-text terms searched in the title, abstract and keyword fields.

No language, publication-type or study-design limits were applied. No animal or human filter was applied, as both preclinical and clinical studies were eligible for inclusion. Both British and American spelling variants were searched throughout.

**Syntax key.** / denotes a controlled vocabulary heading (MeSH in MEDLINE); exp indicates the heading was exploded to retrieve all narrower terms; * denotes truncation, retrieving all variant word endings; adjn (Ovid) to retrieve terms occurring within n words of one another in either order; .ti,ab,kf. searches title, abstract and author keyword fields in MEDLINE.

#### Table S2.1. Ovid MEDLINE(R) ALL

| # | Search terms | Results |
|---|---|---|
| **Concept 1: Haemorrhagic stroke, intracerebral haemorrhage, subarachnoid haemorrhage and intracranial aneurysm** | | |
| 1 | exp Intracranial Hemorrhages/ | 10850 |
| 2 | exp Subarachnoid Hemorrhage/ | 38855 |
| 3 | exp Intracranial Aneurysm/ | 37721 |
| 4 | Hemorrhagic Stroke/ | 9170 |
| 5 | ((intracerebral or intracranial or intraventricular or cerebral or brain or subarachnoid or lobar or putaminal or thalamic) adj3 (haemorrhag* or hemorrhag* or bleed*)).ti,ab,kf. | 10412 |
| 6 | ((haemorrhagic or hemorrhagic) adj2 stroke*).ti,ab,kf. | 20628 |
| 7 | ((intracranial or cerebral or brain or berry or saccular or ruptured) adj3 aneurysm*).ti,ab,kf. | 65189 |
| 8 | (aneurysm* adj3 (ruptur* or bleed* or haemorrhag* or hemorrhag*)).ti,ab,kf. | 10516 |
| 9 | (ICH or SAH or aSAH or IVH).ti,ab,kf. | 55008 |
| 10 | 1 or 2 or 3 or 4 or 5 or 6 or 7 or 8 or 9 | 162072 |
| **Concept 2: Vagus nerve stimulation** | | |
| 11 | Vagus Nerve Stimulation/ | 6142 |
| 12 | Vagus Nerve/ | 32654 |
| 13 | Electric Stimulation/ or Electric Stimulation Therapy/ or Transcutaneous Electric Nerve Stimulation/ | 149492 |
| 14 | 12 and 13 | 17657 |
| 15 | ((vagus or vagal or vagi) adj3 (stimulat* or neurostimulat* or neuromodulat* or electrostimulat*)).ti,ab,kf. | 176576 |
| 16 | (VNS or tVNS or taVNS or nVNS or aVNS or cVNS or tcVNS).ti,ab,kf. | 5262 |
| 17 | ((auricular or auricle or tragus or tragal or cymba or concha* or ear or transauricular or transcutaneous or transcervical or cervical or percutaneous or invasive or noninvasive or non-invasive) adj5 (vagus or vagal)).ti,ab,kf. | 31458 |
| 18 | (gammacore or gamma-core or electrocore or vivistim or microtransponder or nemos or cerbomed or parasym or nurosym or aspireSR or sentiva).ti,ab,kf. | 14699 |
| 19 | ("cholinergic anti-inflammatory pathway" or "cholinergic antiinflammatory pathway").ti,ab,kf. | 995 |

| # | Search terms | Results |
|---|---|---|
| 20 | 11 or 14 or 15 or 16 or 17 or 18 or 19 or 20 | 206088 |
| **Combination of concepts** | | |
| 21 | 10 and 21 | 409 |
